# Integration of a fasting-mimicking diet program in primary care for type 2 diabetes reduces the need for medication – a 12-month randomised controlled trial

**DOI:** 10.1101/2023.10.09.23296744

**Authors:** Elske L van den Burg, Marjolein P Schoonakker, Petra G van Peet, Elske M van den Akker – van Marle, Hildo J Lamb, Valter D Longo, Mattijs E Numans, Hanno Pijl

## Abstract

**Aims/hypothesis:** The aim of this study was to evaluate the impact on metabolic control of the periodic use of a 5-day fasting-mimicking diet (FMD) program as an adjunct to usual care in people with type 2 diabetes under regular primary care surveillance.

**Methods:** In this randomised, controlled, assessor-blinded trial, people with type 2 diabetes using metformin only and/or diet alone for glycaemic control were randomised to receive 5-day cycles of FMD monthly as adjunct to regular care by their general practitioner or regular care only. Primary outcomes were changes in glucose-lowering medication and HbA1c levels after 12 months. Moreover, changes in use of glucose-lowering medication and/or HbA1c levels in individual participants were combined to yield a clinically relevant primary outcome measure (‘glycaemic management’), categorized as improved, stable or deteriorated after one year of follow-up.

**Results:** 100 individuals with type 2 diabetes, age 18-75 years, and BMI > 27 kg/m^2^, were randomised to the FMD (n=51) or control group (n=49). Eight FMD participants and ten controls were lost to follow-up. In complete case intention-to-treat analyses, the mean medication effect score (MES) significantly declined in patients receiving FMD as compared to controls (FMD −0.2 ± 0.3 vs controls +0.2 ± 0.4, p<0.0001) in the face of similar changes of HbA1c adjusted for MES (FMD −0.4 ± 0.8 % vs controls +0.2 ± 0.8 %, p=0.0021). Glycaemic management improved in 53% of participants using FMD vs 8% of controls, remained stable in 23% vs 33%, and deteriorated in 23% vs 59% (p<0.0001).

**Conclusions/interpretation:** Integration of a monthly FMD program in regular primary care for people with type 2 diabetes who use metformin only and/or diet alone for glycaemic control reduces the need for glucose-lowering medication and appears to be safe in routine clinical practice.

**Trial registration:** ClinicalTrials.gov: NCT03811587

## Introduction

Fasting evokes evolutionary conserved adaptive hormonal and cellular responses that enhance stress resistance, dampen inflammation, and optimize metabolism [1]. Experimental studies consistently show robust disease-modifying effects of dietary restriction and intermittent fasting in distinct animal models of chronic disease, including obesity, various cancers, neurodegenerative disorders, and diabetes [2–6]. Various methods of intermittent and periodic energy restriction have shown variable effects on glycaemic control in people with type 2 diabetes[7]. Limiting (formula) dietary intake to 850 kcal/day for 12-20 weeks, followed by structural support for weight loss maintenance, facilitates disease remission in people with type 2 diabetes [8–10]. However, severely restricting calorie intake for extended periods is burdensome for many people and reduces energy expenditure [11], rendering weight maintenance a challenge in the long term [12].

Periodic fasting-mimicking diet (FMD) programs lasting 4 to 7 consecutive days are designed to mimic the physiological effects of water-only fasting while minimizing its burden by allowing patients to consume light meals during the fasting period, while confining it to a limited number of days no more than once a month. These low-calorie plant-based formula diets are low in sugar and protein, primarily comprising complex carbohydrates and healthy plant-based fats. In mice, periodic FMD cycles ameliorate the metabolic anomalies of type 2 diabetes, reverse defects in insulin production [13], and prevent premature death caused by high-fat/high-calorie diets [14]. In healthy (non-diabetic) humans, three 5-day cycles of FMD monthly were shown to reduce fat mass, blood pressure, triglyceride levels, and fasting glucose, particularly in people with high levels of these risk factors at baseline [15].

The vast majority (90%) of people with type 2 diabetes are under primary care surveillance in the Netherlands [16]. Here, we evaluated the clinical response to 5-consecutive-day FMD cycles monthly as an adjunct to regular care in comparison to regular care only in people with type 2 diabetes in a ‘real world’ setting, i.e. under regular primary care surveillance and treatment.

## Methods

### Study design

The study was designed as a randomised, controlled, assessor-blinded intervention trial conducted at the Leiden University Medical Centre (LUMC) in the Netherlands. The trial was performed according to the principles of the Declaration of Helsinki, in accordance with the Medical Research Involving Human Subjects Act (WMO), and to the standards of Good Clinical Practice (GCP). The Medical Research Ethics Committee of the LUMC approved the protocol and amendments. The study was registered as ClinicalTrials.gov NCT03811587, and the study protocol was published [17]. Registration of the trial was initiated prior to the start of the trial; online publication was however realized after the start of the trial due to delay within the registration process.

### Participants

In collaboration with general practice centres, eligible participants under regular primary care surveillance were informed of the study via a letter detailing the trial. Individuals with type 2 diabetes, a BMI≥ 27 kg/m^2^, aged >18 years and <75 years, were eligible. They had to be treated with lifestyle advice only, whilst their glycated haemoglobin (HbA1c) was above 48 mmol/mol (6.5 %) or treated with lifestyle advice plus metformin as the only glucose-lowering drug, irrespective of their HbA1c. A recent myocardial infarction (<6 months), creatinine clearance <30 ml/min/1.73m2, pregnancy, contraindications for MRI, allergy for ingredients of the diet, history of syncope during caloric restriction or any significant other diseases (at the discretion of the investigator) were exclusion criteria. 129 interested individuals were assessed for eligibility; 100 were included after written informed consent.

### Intervention

Participants were allocated to the FMD or control group in computer-generated random sequence via the electronic trial database Castor EDC, which secured allocation concealment. Permuted block randomization was performed with block sizes 2 and 4, stratified for gender and weight <100 kg or >100 kg. Due to its nature, blinding of participants to the intervention was impossible, but study research staff who collected outcome data remained unaware of treatment allocation.

Both the control group and the FMD group received usual care through their general practitioner’s office. Usual care entailed 3-monthly clinical and biochemical evaluation, lifestyle advice with the option to consult a dietitian, and eventual adaptation of medication use according to Dutch guidelines for general practitioners [18]. The study staff did not interfere with usual care in any way. The FMD group received twelve 5-consecutive-day FMD cycles monthly as an adjunct to usual care. Participants were contacted by telephone once during each FMD period to support compliance. The FMD comprised complete meal replacement products (Table S1). Ingredients were all plant-based and generally regarded as safe. Caloric content and macronutrient composition were as follows; day 1 contained ∼ 1100 kcal (10% protein, 56% fat and 34% complex carbohydrate); days 2–5 were identical and provided ∼ 3150 kJ (750 kcal, 9% protein, 44% fat, 47% complex carbohydrate) [17]. The diet of participants who weighed more than 100 kg was supplemented with one bar a day (90 kcal) with similar macronutrient composition. The control group received usual care only. Adherence to the trial regimen was checked verbally every month. We strongly encouraged the participants to complete as many study visits as feasible, even if they decided to quit their assigned treatment, to render missing data as independent of treatment allocation as possible.

### Outcome measures

The participants came to our research unit for baseline- and follow-up visits. HbA1c, total cholesterol, LDL-cholesterol, HDL-cholesterol, triglycerides, and high-sensitive CRP were measured in fasting condition at baseline and after 6 and 12 months. Plasma glucose and insulin concentrations were measured several times over the course of 2 hours during the oral glucose tolerance test (OGTT). Bodyweight, waist circumference, body fat percentage, and blood pressure were measured every visit. All measurements at 6 and 12 months were performed three weeks after the last FMD cycle in those who received FMD.

The primary outcomes were change of HbA1c and dosage of glucose-lowering medication from baseline. Mean change in glucose-lowering medication use was quantified by use of the medication effect score (MES). The MES reflects the overall intensity of a glucose-lowering medication regimen based on medication dosages and their potential efficacy in terms of reducing blood glucose [19]. The MES was calculated for each diabetes drug in a regimen using the following equation: (actual drug dose/maximum drug dose) × drug-specific adjustment factor. The adjustment factor corresponds to the expected decrease in HbA1c achieved by the drug as monotherapy. The sum of MES values attributed to individual drugs represents the maximum HbA1c reduction that may be expected by the regimen [19]. Additionally, HbA1c was corrected for MES by calculating the sum of MES values and HbA1c levels.

As the response of individual participants (in addition to average results) provides valuable insight into the clinical effects of an intervention, we also categorized both outcome measures in each individual participant. The categories are described in Table 1. As plasma HbA1c concentration and the dose of glucose-lowering drugs mutually influence each other, we combined these parameters reflecting glucose control in individual participants to yield a categorical outcome measure, for which we coined the term ‘glycaemic management’ (Table 1).

**Table 1.** Categories of individual change at the end of the study compared to baseline for HbA1c, glucose-lowering medication use and glycaemic management.

| **HbA1c levels** |  |
| --- | --- |
| Improved | ≥ 5 mmol/mol (0.5 %) lower |
| Stable | < 5 mmol/mol (0.5 %) higher or lower: stable |
| Deteriorated | ≥ 5 mmol/mol (0.5 %) higher |
| **Glucose-lowering medication use** |  |
| Decreased | lower dose or stop metformin |
| Stable | stable dose |
| Increased | increased dose of metformin and/or additional drugs to control glycemia |
| **Glycaemic management (change in HbA1c levels and use of glucose-lowering medication combined)** |  |
| Improved | a lower dose or class of glucose-lowering medication with an HbA1c not more than 5 mmol/mol (0.5 %) higher at the end of the study compared to baseline or;<br>no change in glucose-lowering medication with an HbA1c that is ≥ 5 mmol/mol (0.5 %) lower at the end of the study compared to baseline |
| Stable | no change in glucose-lowering medication use and a difference in HbA1c of < 5 mmol/mol (0.5 %) at the end of the study compared to baseline |
| Deteriorated | a higher dose or class of glucose-lowering medication at the end of the study compared to baseline or;<br>an HbA1c that is ≥ 5 mmol/mol (0.5 %) higher at the end of the study compared to baseline with no change in glucose-lowering medication |

Secondary outcomes were body weight, BMI, total body fat, waist circumference, blood pressure, fasting plasma glucose, insulin, and lipid profiles. Furthermore, plasma glucose and insulin concentrations in response to an oral glucose tolerance test were used to calculate the Matsuda index (reflecting insulin sensitivity) and the disposition index (reflecting endogenous insulin secretion) [20–22]. Adverse events were registered according to the Common Terminology Criteria for Adverse Events version 5.0 [23] during two face-to-face visits at 6 and 12 months or (in case of serious adverse events) reported immediately.

### Statistical analysis

Assuming glycaemic control to improve in 5% of controls [9, 24], inclusion of 45 participants in each group would yield 80% power to detect an absolute difference with FMD of at least 21% (i.e., improvement in 5% of controls vs 26% of FMD) at a significance level of 5% using a two-sided binomial test.

Primary and secondary outcomes were summarized using the mean and standard deviation or median and interquartile range in case of an asymmetric distribution. The categorical primary outcome measures were analysed using chi-square tests. When the assumptions of the chi-square test were violated, the Fischer’s exact test was used. Average HbA1c and secondary outcomes were compared between the two groups using independent t-tests or a Mann-Whitney U test if the assumption of normality was violated with a confidence level of 95%. The procedure of Benjamini-Hochberg was used to correct the statistics of the multiple tests of secondary outcomes. A complete case intention-to-treat (ITT) analysis was conducted as well as a per protocol (PP) analysis, including only participants in the FMD group who were compliant with the 12 cycles of FMD. Imputation was not performed, for this could only be applied to the outcome measure, where no power or efficiency would be gained. Last measurement carried forward was not applied because of the bias it would introduce [25].

Additionally, changes over time in HbA1c, MES and HbA1c corrected for MES were estimated with linear mixed models using all available data at baseline, 6 months and 12 months (ITT). The outcome model included fixed effects for time-by-arm interaction terms with random effects for individual participants. As a post-hoc analysis, we added the interaction between weight and intervention group as a fixed effect to the linear mixed models. Statistical analyses were done using Rstudio version 4.1.0 for Windows. Figures were created in GraphPad Prism version 9.0.1 for Windows.

## Results

### Trial participants

Between November 20, 2018, and July 1, 2020, 129 individuals were assessed for eligibility, of whom 29 were excluded; 100 participants were randomly assigned to the FMD- or control group (FMD n=51; control n=49, Fig. 1). Follow-up ended on August 5, 2021.

**Figure 1:**
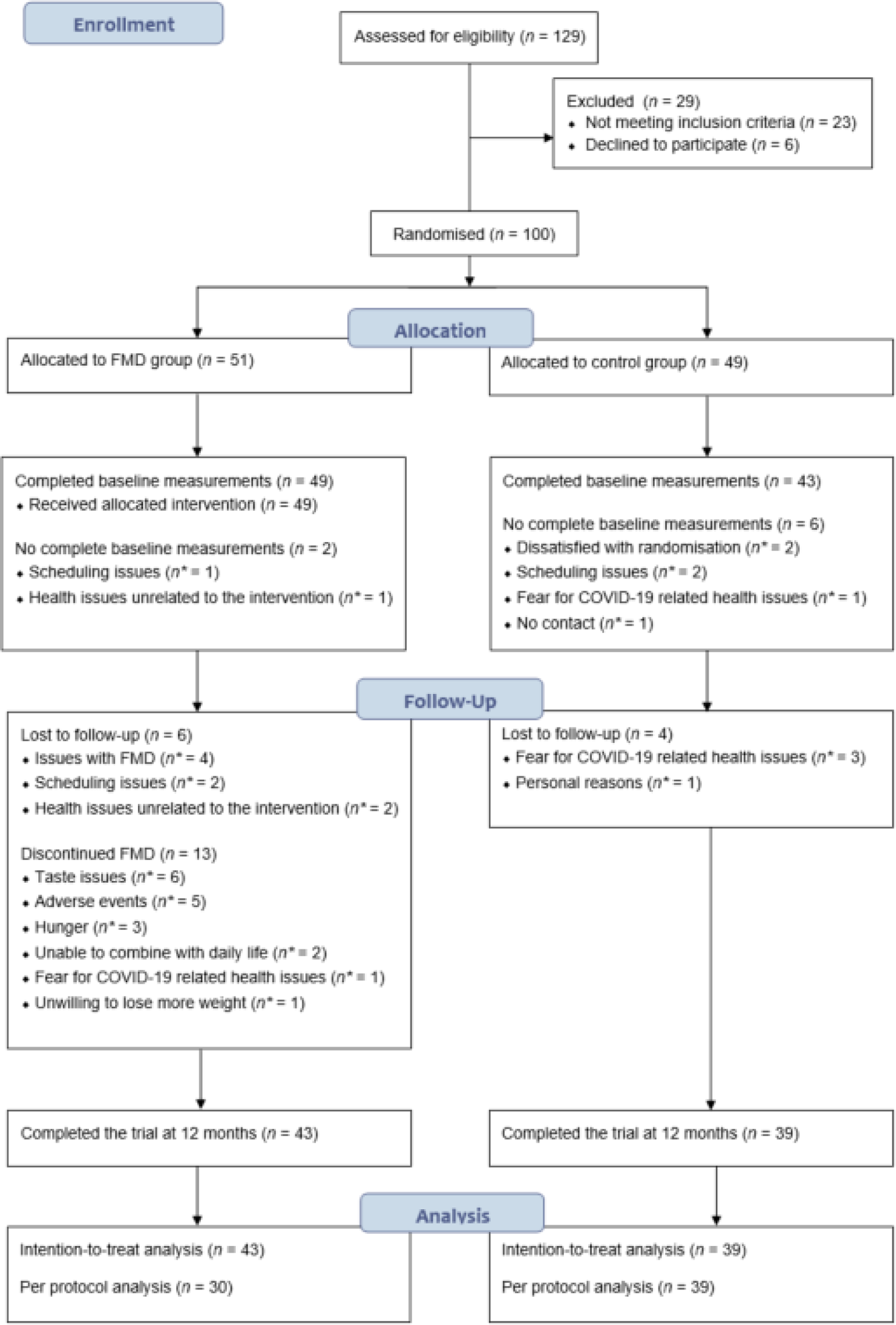
Study flow chart. COVID-19 = coronavirus disease 2019. FMD = fasting-mimicking diet. n = number of participants. n* = number of participants for whom this was reason for being lost to follow-up or discontinuing FMD, there may be several reasons per participant.

Two participants in the FMD group and six participants in the control group did not complete baseline measurements. Furthermore, despite strong encouragement, six non-compliant FMD participants and four controls could not complete follow-up visits. Indeed, lost-to-follow-up was primarily due to the inability to complete study visits and unrelated to treatment issues. Moreover, lost-to-follow-up participants were equally distributed among study groups. For these reasons, missing data were assumed to be random. At various time points during the protocol, 13 other participants stopped using the FMD, but agreed to complete follow-up visits. Thus, the data of 43 participants using FMD and 39 controls could be used for the intention-to-treat analysis (Fig. 1). Demographics and clinical characteristics were similar in both groups at baseline (Table 2). On average, glucose metabolism was well controlled, as indicated by an on-target HbA1c.

**Table 2.**
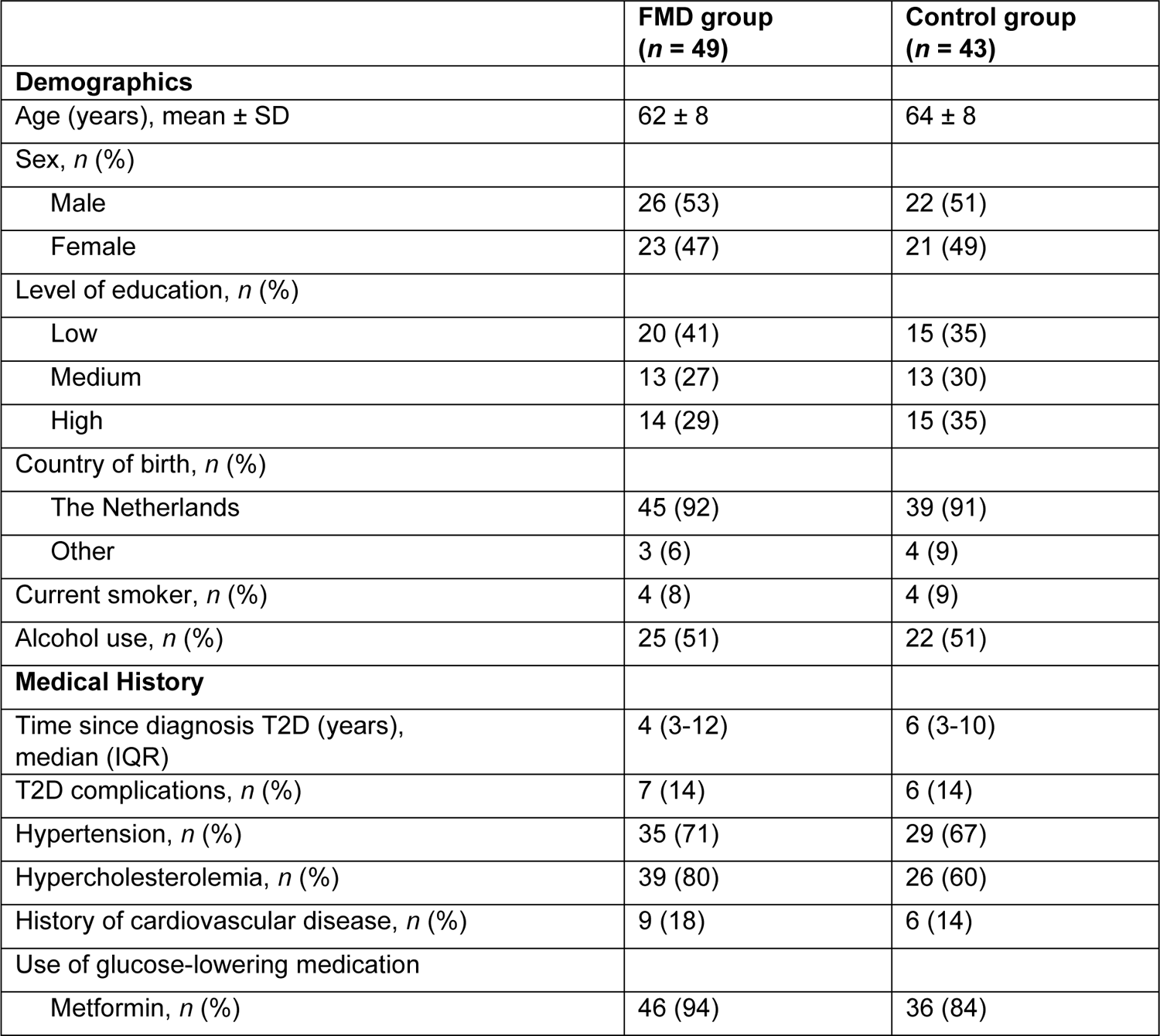
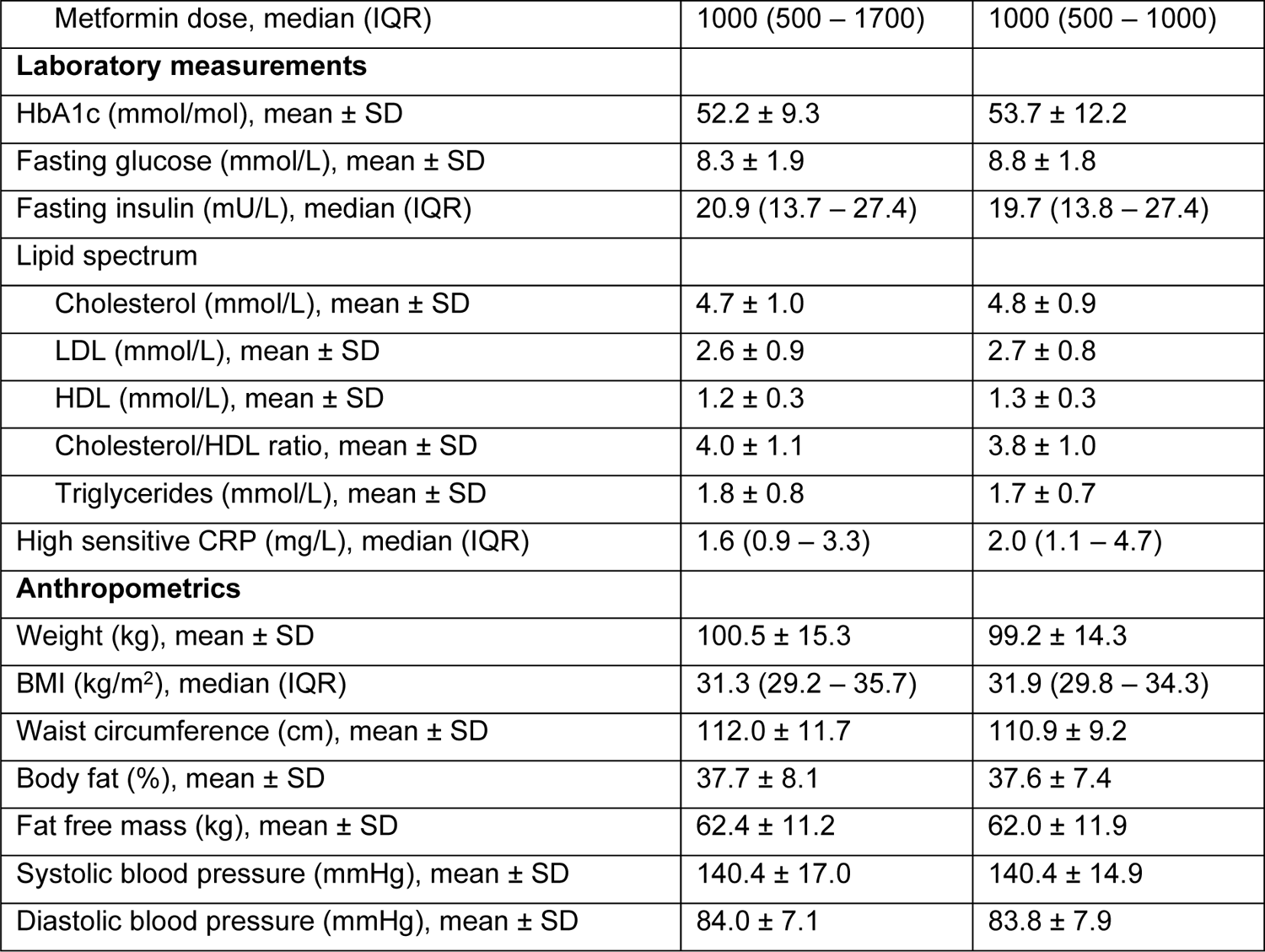
Demographics and baseline characteristics (n=92) Data are presented as mean ± SD, median (IQR) or number (*n*) with percentage (%). T2D complications include polyneuropathy and retinopathy. There were no cases of nephropathy or diabetic foot. History of cardiovascular disease includes angina pectoris, myocardial infarction, and cerebrovascular events. Missings: One patient in each group did not arrive in fasting condition, therefore fasting glucose and fasting insulin are missing. In the FMD group, one fasting insulin measurement was invalid. The plasma lipid spectrum (except HDL) is missing for one patient in the control group due to invalid measurement. Two patients in the FMD group refused to share their level of education, one patient in the FMD group refused to share information on country of birth. BMI=Body Mass Index. CRP=c-reactive protein. FMD=fasting-mimicking diet. HbA1c=glycated haemoglobin. T2D=type 2 diabetes.

|  | **FMD group**<br>**(n = 49)** | **Control group**<br>**(n = 43)** |
| --- | --- | --- |
| **Demographics** |  |  |
| Age (years), mean ± SD | 62 ± 8 | 64 ± 8 |
| Sex, n (%) |  |  |
| Male | 26 (53) | 22 (51) |
| Female | 23 (47) | 21 (49) |
| Level of education, n (%) |  |  |
| Low | 20 (41) | 15 (35) |
| Medium | 13 (27) | 13 (30) |
| High | 14 (29) | 15 (35) |
| Country of birth, n (%) |  |  |
| The Netherlands | 45 (92) | 39 (91) |
| Other | 3 (6) | 4 (9) |
| Current smoker, n (%) | 4 (8) | 4 (9) |
| Alcohol use, n (%) | 25 (51) | 22 (51) |
| **Medical History** |  |  |
| Time since diagnosis T2D (years), median (IQR) | 4 (3-12) | 6 (3-10) |
| T2D complications, n (%) | 7 (14) | 6 (14) |
| Hypertension, n (%) | 35 (71) | 29 (67) |
| Hypercholesterolemia, n (%) | 39 (80) | 26 (60) |
| History of cardiovascular disease, n (%) | 9 (18) | 6 (14) |
| Use of glucose-lowering medication |  |  |
| Metformin, n (%) | 46 (94) | 36 (84) |
| Metformin dose, median (IQR) | 1000 (500 – 1700) | 1000 (500 – 1000) |
| **Laboratory measurements** |  |  |
| HbA1c (mmol/mol), mean ± SD | 52.2 ± 9.3 | 53.7 ± 12.2 |
| Fasting glucose (mmol/L), mean ± SD | 8.3 ± 1.9 | 8.8 ± 1.8 |
| Fasting insulin (mU/L), median (IQR) | 20.9 (13.7 – 27.4) | 19.7 (13.8 – 27.4) |
| **Lipid spectrum** |  |  |
| Cholesterol (mmol/L), mean ± SD | 4.7 ± 1.0 | 4.8 ± 0.9 |
| LDL (mmol/L), mean ± SD | 2.6 ± 0.9 | 2.7 ± 0.8 |
| HDL (mmol/L), mean ± SD | 1.2 ± 0.3 | 1.3 ± 0.3 |
| Cholesterol/HDL ratio, mean ± SD | 4.0 ± 1.1 | 3.8 ± 1.0 |
| Triglycerides (mmol/L), mean ± SD | 1.8 ± 0.8 | 1.7 ± 0.7 |
| High sensitive CRP (mg/L), median (IQR) | 1.6 (0.9 – 3.3) | 2.0 (1.1 – 4.7) |
| **Anthropometrics** |  |  |
| Weight (kg), mean ± SD | 100.5 ± 15.3 | 99.2 ± 14.3 |
| BMI (kg/m<sup>2</sup>), median (IQR) | 31.3 (29.2 – 35.7) | 31.9 (29.8 – 34.3) |
| Waist circumference (cm), mean ± SD | 112.0 ± 11.7 | 110.9 ± 9.2 |
| Body fat (%), mean ± SD | 37.7 ± 8.1 | 37.6 ± 7.4 |
| Fat free mass (kg), mean ± SD | 62.4 ± 11.2 | 62.0 ± 11.9 |
| Systolic blood pressure (mmHg), mean ± SD | 140.4 ± 17.0 | 140.4 ± 14.9 |
| Diastolic blood pressure (mmHg), mean ± SD | 84.0 ± 7.1 | 83.8 ± 7.9 |

### Glycaemic endpoints

#### Glucose-lowering medication

In the complete case ITT analysis using an independent t-test, the use of glucose-lowering medication after 12 months as quantified by the MES declined more in participants using the FMD (−0.2 ± 0.3, n=42) versus control (+0.2 ± 0.4, n=38, p<0.0001, Fig. 2). Linear mixed models analysis yielded similar results (Supplementary Table S2a). There was no significant effect of the interaction between weight and intervention on the MES (Supplementary Table S2b). The dose of glucose-lowering medication was reduced in 40% (n=17) of participants in the FMD group and in 5% (n=2) of controls, remained stable in 51% (n= 22) of participants receiving FMD and in 51% (n=20) of controls, and increased in 9% (n=4) of participants using FMD and 44% (n=17) of controls (p<0.001, Fig. 3). Glucose-lowering medication was completely stopped in 16% (n=7) of the participants in the FMD group and in 5% (n=2) of controls (p=0.16), while additional medication was prescribed in 2.3% (n=1) of the FMD group and 25.6% (n=10) of the control group (p=0.006, Fig 4).

**Figure 2:**
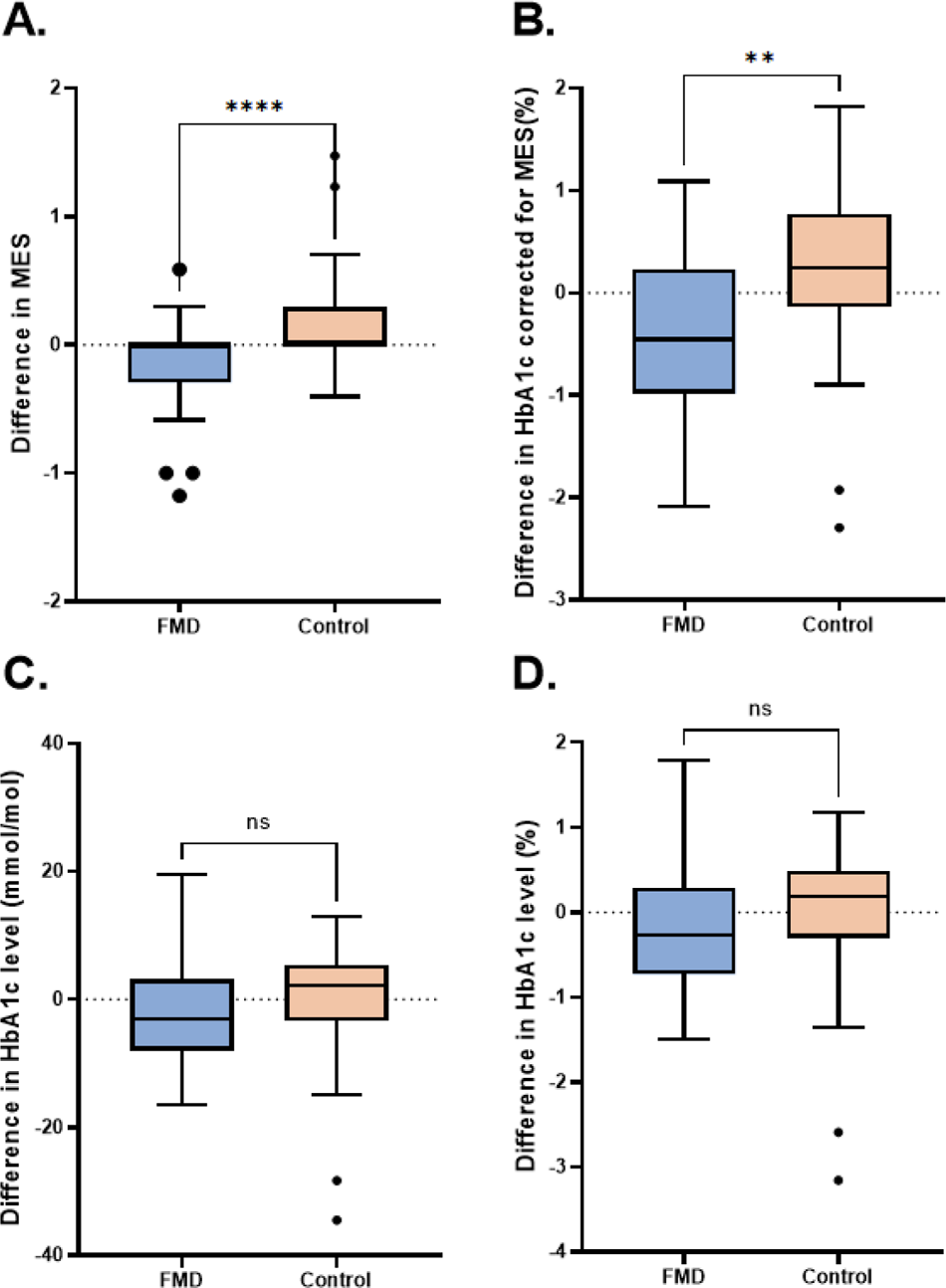
Average change in MES, HbA1c and HbA1c corrected for MES from baseline to 12 months in the FMD and control group (intention-to-treat analysis) The box-and-whisker plot with the Tukey method for whiskers and extreme outliers represents the change from baseline to end of intervention. Differences between FMD- and control group were evaluated using an independent t-test. Number of participants with data available at baseline and 12 months were used for each outcome. (A) Change in medication effect score (MES), n=42 in the FMD group vs n = 38 in the control group. (B) Change in HbA1c corrected for MES (%), n = 42 in the FMD group vs n = 38 in the control group (C) Change in HbA1c (mmol/mol), n=43 in the FMD group vs n = 39 in the control group (D) Change in HbA1c (%), n=43 in the FMD group vs n = 39 in the control group FMD=fasting-mimicking diet. HbA1c=glycated haemoglobin.

**Figure 3:**
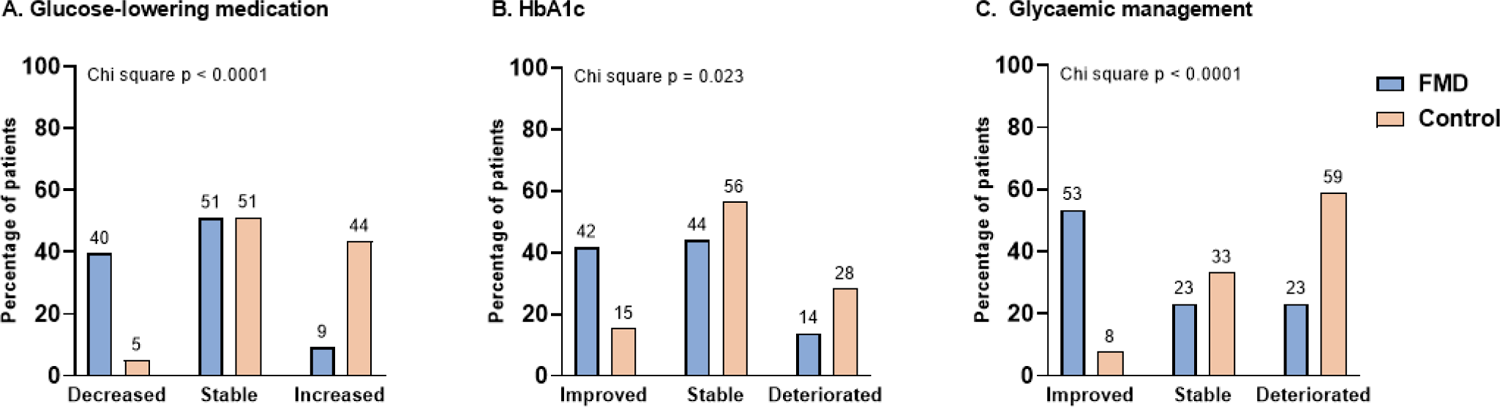
Effect of FMD on glucose-lowering medication use and HbA1c levels in individual participants in the FMD group and the control group at 12 months (intention-to-treat analysis) Plotted bars represent percentage of participants (n=43 in the FMD group vs n=39 in the control group). Differences between FMD- and control group were evaluated using the Chi square test. A) Change in glucose-lowering medication, with decrease defined as a lower dose, stable as no change and increased as higher dose or other class of glucose-lowering medication at the end of the study compared to baseline. (B) Change in HbA1c, defined as follows; Improved: an HbA1c that is ≥5 mmol/mol (≥0.5 %) lower at the end of the study compared to baseline. Stable: a change in HbA1c of <5 mmol/mol (<0.5 %) at the end of the study compared to baseline. Deteriorated: an HbA1c that is ≥5 mmol/mol (≥0.5 %) higher at the end of the study compared to baseline. (C) Glycaemic management, defined as follows; Improved: a lower dose or class of glucose-lowering medication with an HbA1c not more than 5 mmol/mol (0.5 %) higher at the end of the study compared to baseline or; no change in glucose-lowering medication with an HbA1c that is ≥5 mmol/mol (≥0.5 %) lower at the end of the study compared to baseline. Stable: no change in glucose-lowering medication use and a difference in HbA1c of <5 mmol/mol (<0.5 %) at the end of the study compared to baseline. Deteriorated: a higher dose or class of glucose-lowering medication at the end of the study compared to baseline or; an HbA1c that is ≥5 mmol/mol (≥0.5 %) higher at the end of the study compared to baseline with no change in glucose-lowering medication (Table 1). FMD=fasting-mimicking diet. HbA1c=glycated haemoglobin. Between-group p-values were calculated using chi-square tests.

**Figure 4:**
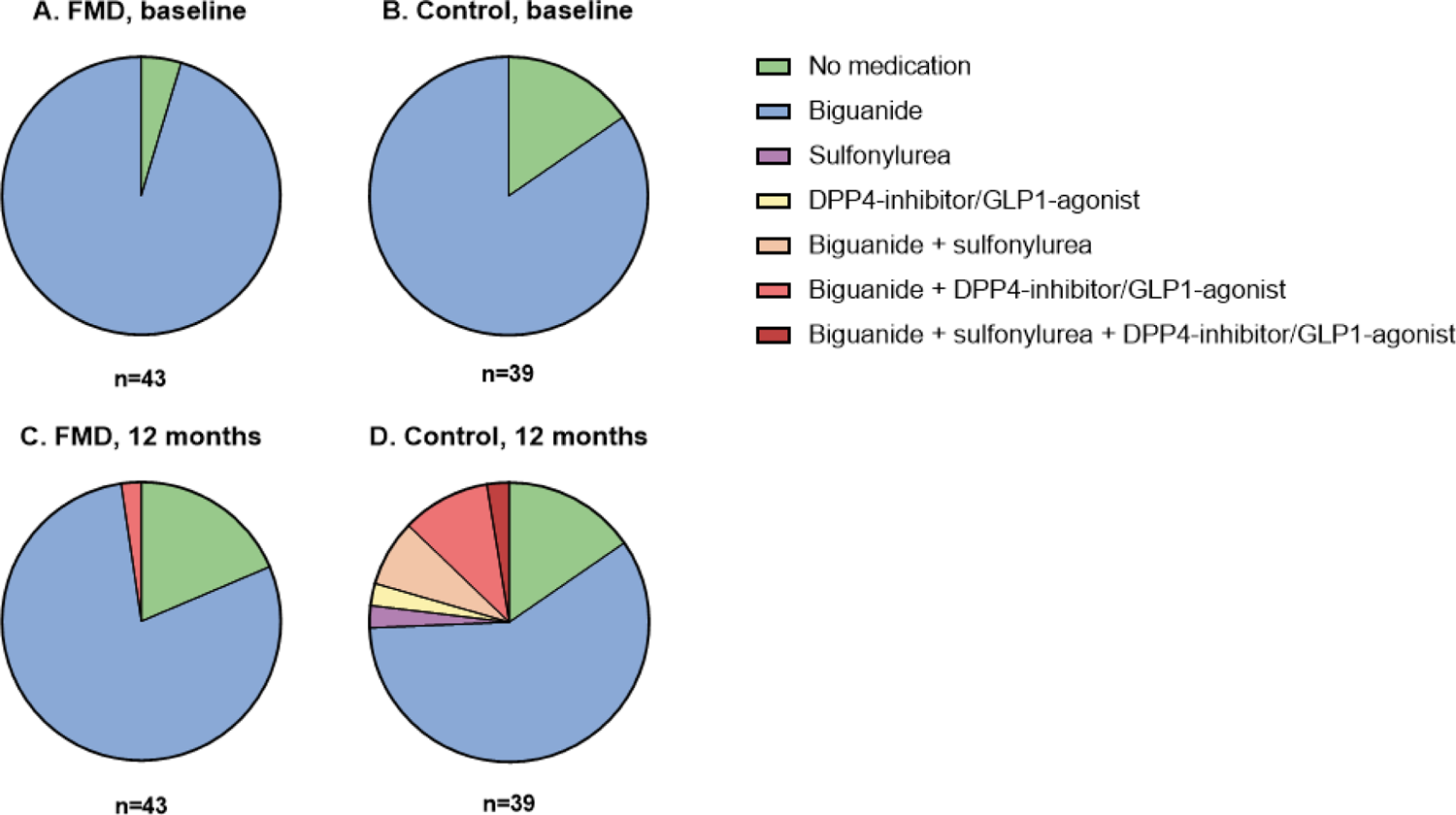
Overview of glucose-lowering medication at baseline and at 12 months in the FMD group and the control group (intention-to-treat analysis) (A) Use of glucose-lowering medication in the FMD group at baseline. (B) Use of glucose-lowering medication in the control group at baseline. (C) Use of glucose-lowering medication in the FMD group after 12 months. (D). Use of glucose-lowering medication the control group after 12 months. DPP4-inhibitor=dipeptidyl-peptidase-4 inhibitor. FMD=fasting-mimicking diet. GLP-1-agonist=glucagon-like peptide-1 agonist.

#### HbA1c

While the use of glucose-lowering medication declined more in the FMD group, the average change of HbA1c did not differ between the groups in the complete case ITT analysis using an independent t-test (FMD: −2.4 ± 8.0 mmol/mol (−0.2 ± 0.7%), n=43, versus control: 0.0 ± 9.6 mmol/mol (0.0 ± 0.9 %), n=38, p=0.22) (Table 3, Fig. 2). Linear mixed models analysis yielded similar results (Supplementary Table S2a). There was no significant effect of the interaction between weight and intervention on HbA1c (Supplementary Table S2b). HbA1c was reduced by >5 mmol in 42% (n=18) of participants in the FMD group and in 15% (n=6) of controls, whereas it remained stable in 44% (n=19) of patients in the FMD group and 56% (n=22) of controls and deteriorated in 14% (n=6) of FMD participants and in 28% (n=11) of controls (p=0.023, Fig 3).

**Table 3.**
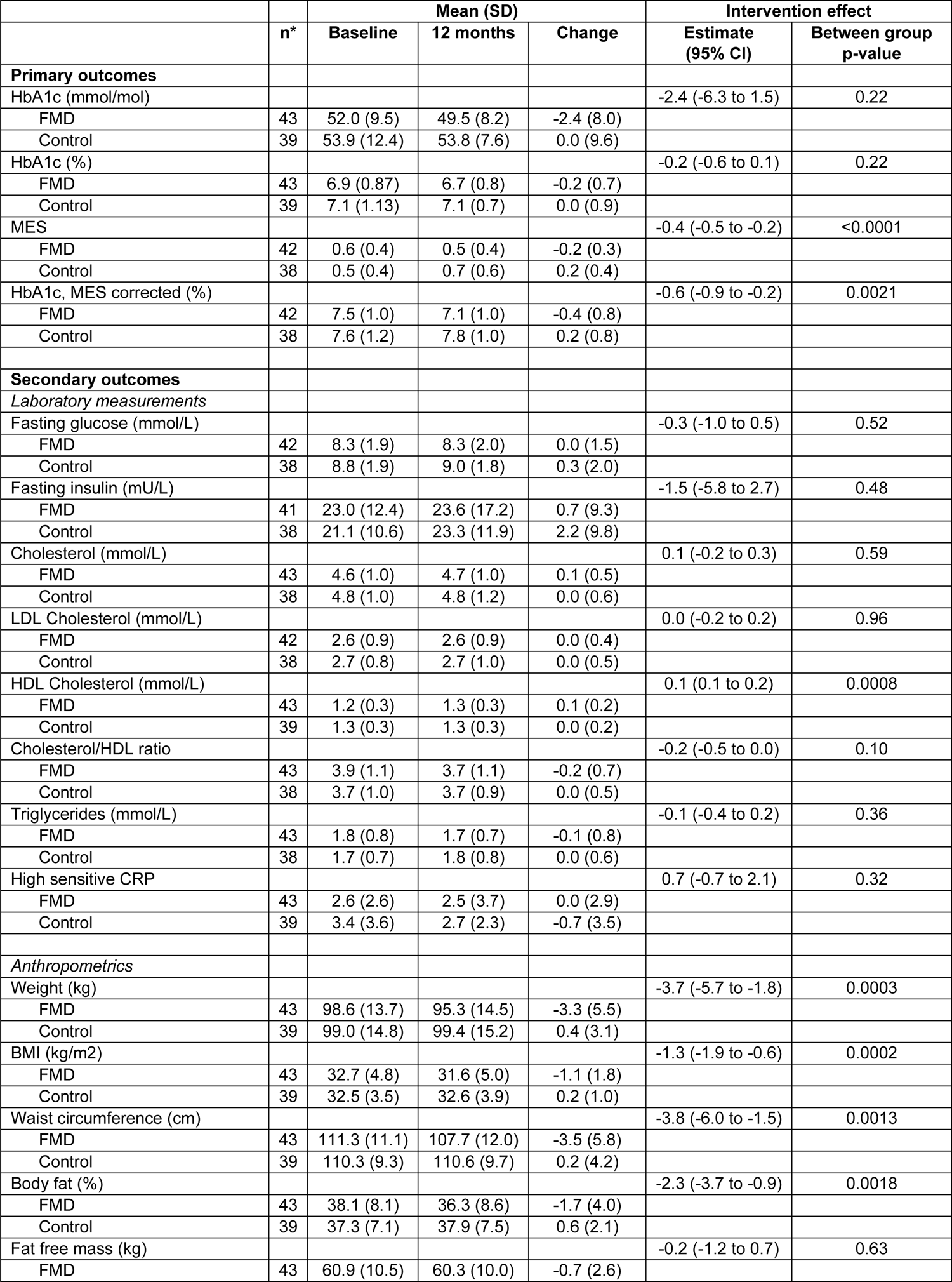
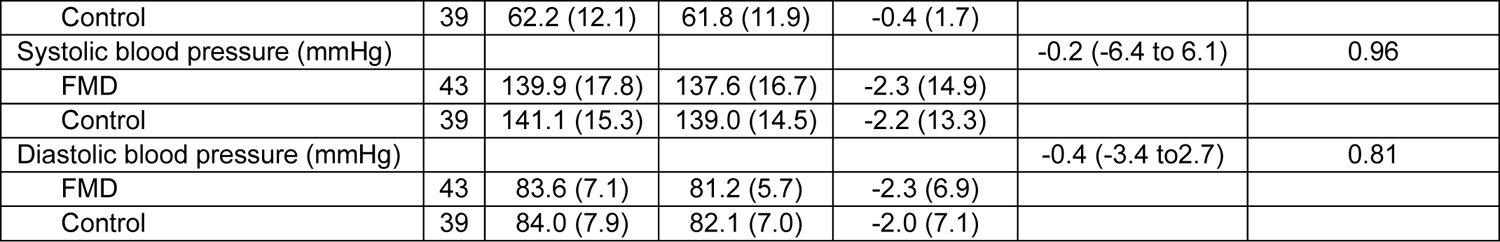
Average change of anthropometrics and plasma metabolic profiles from baseline to end of study in FMD and control group (intention-to-treat analysis) *Number of participants with data available at baseline and 12 months for each outcome. BMI=body mass index. CI=confidence interval. CRP=C-reactive protein. FMD=fasting-mimicking diet. HbA1c=glycated haemoglobin. Between-group p-values were calculated using independent t-tests.

|  |  | Mean (SD) |  |  | Intervention effect |  |
| --- | --- | --- | --- | --- | --- | --- |
|  | n* | Baseline | 12 months | Change | Estimate (95% CI) | Between group p-value |
| **Primary outcomes** |  |  |  |  |  |  |
| HbA1c (mmol/mol) |  |  |  |  | -2.4 (-6.3 to 1.5) | 0.22 |
| FMD | 43 | 52.0 (9.5) | 49.5 (8.2) | -2.4 (8.0) |  |  |
| Control | 39 | 53.9 (12.4) | 53.8 (7.6) | 0.0 (9.6) |  |  |
| HbA1c (%) |  |  |  |  | -0.2 (-0.6 to 0.1) | 0.22 |
| FMD | 43 | 6.9 (0.87) | 6.7 (0.8) | -0.2 (0.7) |  |  |
| Control | 39 | 7.1 (1.13) | 7.1 (0.7) | 0.0 (0.9) |  |  |
| MES |  |  |  |  | -0.4 (-0.5 to -0.2) | <0.0001 |
| FMD | 42 | 0.6 (0.4) | 0.5 (0.4) | -0.2 (0.3) |  |  |
| Control | 38 | 0.5 (0.4) | 0.7 (0.6) | 0.2 (0.4) |  |  |
| HbA1c, MES corrected (%) |  |  |  |  | -0.6 (-0.9 to -0.2) | 0.0021 |
| FMD | 42 | 7.5 (1.0) | 7.1 (1.0) | -0.4 (0.8) |  |  |
| Control | 38 | 7.6 (1.2) | 7.8 (1.0) | 0.2 (0.8) |  |  |
| **Secondary outcomes** |  |  |  |  |  |  |
| *Laboratory measurements* |  |  |  |  |  |  |
| Fasting glucose (mmol/L) |  |  |  |  | -0.3 (-1.0 to 0.5) | 0.52 |
| FMD | 42 | 8.3 (1.9) | 8.3 (2.0) | 0.0 (1.5) |  |  |
| Control | 38 | 8.8 (1.9) | 9.0 (1.8) | 0.3 (2.0) |  |  |
| Fasting insulin (mU/L) |  |  |  |  | -1.5 (-5.8 to 2.7) | 0.48 |
| FMD | 41 | 23.0 (12.4) | 23.6 (17.2) | 0.7 (9.3) |  |  |
| Control | 38 | 21.1 (10.6) | 23.3 (11.9) | 2.2 (9.8) |  |  |
| Cholesterol (mmol/L) |  |  |  |  | 0.1 (-0.2 to 0.3) | 0.59 |
| FMD | 43 | 4.6 (1.0) | 4.7 (1.0) | 0.1 (0.5) |  |  |
| Control | 38 | 4.8 (1.0) | 4.8 (1.2) | 0.0 (0.6) |  |  |
| LDL Cholesterol (mmol/L) |  |  |  |  | 0.0 (-0.2 to 0.2) | 0.96 |
| FMD | 42 | 2.6 (0.9) | 2.6 (0.9) | 0.0 (0.4) |  |  |
| Control | 38 | 2.7 (0.8) | 2.7 (1.0) | 0.0 (0.5) |  |  |
| HDL Cholesterol (mmol/L) |  |  |  |  | 0.1 (0.1 to 0.2) | 0.0008 |
| FMD | 43 | 1.2 (0.3) | 1.3 (0.3) | 0.1 (0.2) |  |  |
| Control | 39 | 1.3 (0.3) | 1.3 (0.3) | 0.0 (0.2) |  |  |
| Cholesterol/HDL ratio |  |  |  |  | -0.2 (-0.5 to 0.0) | 0.10 |
| FMD | 43 | 3.9 (1.1) | 3.7 (1.1) | -0.2 (0.7) |  |  |
| Control | 38 | 3.7 (1.0) | 3.7 (0.9) | 0.0 (0.5) |  |  |
| Triglycerides (mmol/L) |  |  |  |  | -0.1 (-0.4 to 0.2) | 0.36 |
| FMD | 43 | 1.8 (0.8) | 1.7 (0.7) | -0.1 (0.8) |  |  |
| Control | 38 | 1.7 (0.7) | 1.8 (0.8) | 0.0 (0.6) |  |  |
| High sensitive CRP |  |  |  |  | 0.7 (-0.7 to 2.1) | 0.32 |
| FMD | 43 | 2.6 (2.6) | 2.5 (3.7) | 0.0 (2.9) |  |  |
| Control | 39 | 3.4 (3.6) | 2.7 (2.3) | -0.7 (3.5) |  |  |
| *Anthropometrics* |  |  |  |  |  |  |
| Weight (kg) |  |  |  |  | -3.7 (-5.7 to -1.8) | 0.0003 |
| FMD | 43 | 98.6 (13.7) | 95.3 (14.5) | -3.3 (5.5) |  |  |
| Control | 39 | 99.0 (14.8) | 99.4 (15.2) | 0.4 (3.1) |  |  |
| BMI (kg/m2) |  |  |  |  | -1.3 (-1.9 to -0.6) | 0.0002 |
| FMD | 43 | 32.7 (4.8) | 31.6 (5.0) | -1.1 (1.8) |  |  |
| Control | 39 | 32.5 (3.5) | 32.6 (3.9) | 0.2 (1.0) |  |  |
| Waist circumference (cm) |  |  |  |  | -3.8 (-6.0 to -1.5) | 0.0013 |
| FMD | 43 | 111.3 (11.1) | 107.7 (12.0) | -3.5 (5.8) |  |  |
| Control | 39 | 110.3 (9.3) | 110.6 (9.7) | 0.2 (4.2) |  |  |
| Body fat (%) |  |  |  |  | -2.3 (-3.7 to -0.9) | 0.0018 |
| FMD | 43 | 38.1 (8.1) | 36.3 (8.6) | -1.7 (4.0) |  |  |
| Control | 39 | 37.3 (7.1) | 37.9 (7.5) | 0.6 (2.1) |  |  |
| Fat free mass (kg) |  |  |  |  | -0.2 (-1.2 to 0.7) | 0.63 |
| FMD | 43 | 60.9 (10.5) | 60.3 (10.0) | -0.7 (2.6) |  |  |
| Control | 39 | 62.2 (12.1) | 61.8 (11.9) | -0.4 (1.7) |  |  |
| Systolic blood pressure (mmHg) |  |  |  |  | -0.2 (-6.4 to 6.1) | 0.96 |
| FMD | 43 | 139.9 (17.8) | 137.6 (16.7) | -2.3 (14.9) |  |  |
| Control | 39 | 141.1 (15.3) | 139.0 (14.5) | -2.2 (13.3) |  |  |
| Diastolic blood pressure (mmHg) |  |  |  |  | -0.4 (-3.4 to 2.7) | 0.81 |
| FMD | 43 | 83.6 (7.1) | 81.2 (5.7) | -2.3 (6.9) |  |  |
| Control | 39 | 84.0 (7.9) | 82.1 (7.0) | -2.0 (7.1) |  |  |

### Glucose-lowering medication and HbA1c combined

Since our primary outcome measures were both glucose-lowering medication use and HbA1c, these measures were combined to indicate clinical improvement of the type 2 diabetes status. When change in HbA1c was adjusted for the medication effect score (MES), groups did differ in the complete case ITT analysis using an independent t-test, as mean HbA1c declined in the FMD group, whereas it increased in the control group (−0.4 ± 0.8 %, n=42 versus +0.2 ± 0.8 %, n=38, p=0.0021, Fig. 2). Linear mixed models analysis yielded similar results (Supplementary Table S2a). There was no significant effect of the interaction between weight and intervention on HbA1c adjusted for the MES (Supplementary Table S2b).

Furthermore, in our analyses of the outcome measure ‘glycaemic management’ (Table 1, Fig. 3), 53% (n=23) of individual participants of the FMD group improved compared to 8% (n=3) of controls, while 23% (n=10) of patients receiving FMD and 33% (n=13) of controls remained stable, and 23% (n=10) using FMD and 59% (n=23) of controls deteriorated (p<0.0001). Two participants in the FMD group could not formally be categorized, since they had a rise in HbA1c of > 5 mmol/mol while they used less glucose-lowering medication after 12 months. We subjectively decided to categorize these patients as ‘deteriorated’.

### Matsuda and disposition index

The Matsuda index and the disposition index were calculated using glucose and insulin data obtained during an OGTT (Supplementary Fig. S1). The mean change in Matsuda index from baseline to 12 months differed between groups (+0.30 ± 0.85 (n=31) in the FMD group and −0.15 ± 0.46 (n=35) in the control group, p=0.012). The change in disposition index from baseline to 12 months did not differ between groups (mean difference 1.83, 95% CI −6.07 to +2.41, p=0.39).

### Per protocol analysis

For the per protocol (PP) analysis, data from 30 participants who were fully compliant with the dietary program were compared with data from the control group (n=39, same as ITT) (Supplementary Fig. S2 and table S3). Mean values of MES and HbA1c changed to a similar extent in PP and ITT analyses. Medication use decreased in 47% (n=14) of participants in the FMD group and in 5% (n=2) of controls, remained stable in 47% (n=14) of participants in the FMD group and in 51% (n=20), and increased in 7% (n=2) of FMD compliant participants and 44% (n=17) of controls (p < 0.0001). HbA1c improved in 50% (n=15) of participants in the FMD group and in 15% (n=6) of controls, whereas it remained stable in 37% (n=11) of participants in the FMD group and 56% (n=22) of controls and deteriorated in 13% (n=4) of FMD participants and in 28% (n=11) of controls (p=0.0075). Glycaemic management improved in 63% (n=19) of FMD compliant participants compared to 8% (n=3) of controls, whereas it remained stable in 17% (n=5) of FMD compliant participants compared to 33% (n=13) of controls and deteriorated in 20% (n=6) of compliant participants using FMD and 59% (n=23) of controls (p<0.0001).

### Anthropometrics and plasma lipid profiles

Bodyweight (−3.7kg, 95% CI −5.7 to −1.8, p<0.001), BMI (−1.3 kg/m^2^, 95% CI −1.9 to −0.6, p<0.001), waist circumference (−3.8cm, 95% CI −6.0 to −1.5, p=0.0013) and body fat percentage (−2.3%, 95% CI −3.7 to −0.9, p=0.0018) declined more in participants receiving FMD than in controls after 12 months, while the change in fat free mass did not differ between the FMD- and control group (−0.2 kg, 95% CI −1.2 to 0.7, p=0.63)(Table 2). Also, changes in systolic blood pressure (SBP) and diastolic blood pressure (DBP) did not differ (SBP −0.2mmHg, 95% CI −6.4 to +6.1, p=0.96; DBP −0.4mmHg, 95% CI −3.4 to +2.7, p=0.81, Table 2), in the face of largely unchanged antihypertensive drug use (63% of participants receiving FMD and 79% of controls used similar antihypertensive medication after 12 months).

The change in plasma lipids did not differ between groups, except for the HDL-cholesterol concentration, which increased only in the FMD group (+0.1mmol/L, 95% CI +0.1 to +0.2, p<0.001, Table 2). Use of cholesterol lowering medication remained stable over 12 months in the vast majority of participants (80% of participants receiving FMD versus 84% of controls). All parameters significantly different between groups remained so after correction for multiple testing by the Benjamini-Hochberg procedure.

Notably, although the primary outcome measures, as well as anthropometrics and plasma metabolic profiles tended to improve to a maximum extent of 6 months, the MES appeared to decline further over the next 6 months of intervention (Fig. S4, Table S4).

### Adverse events

FMD caused typical signs of energy deficit (fatigue, headache, dizziness) and nausea in a substantial number of patients during the 5-day intervention, which resolved in the periods between the FMD cycles. Adverse events were registered in 19 FMD and 18 control participants (Table S5). Eight serious adverse events occurred; none were related to the study (Table S6).

## Discussion

We explored the clinical impact of periodic use of an FMD program as adjunct to usual care for people with type 2 diabetes. The data show that, on average, the group assigned to 12 cycles of 5-consecutive days of FMD monthly used significantly less glucose-lowering medication, while HbA1c levels remained similar to those in the control group. Indeed, the proportion of participants who could reduce glucose-lowering medication was 8 times higher in the FMD − (40%) than in the control group (5%). Interestingly, although the change of mean HbA1c levels did not differ between groups, HbA1c declined ≥ 5 mmol/mol (0.5 %) in 42% of individual participants in the FMD group, while this happened in only 15% of participants in the control group. Moreover, average bodyweight, body fat percentage and waist circumference declined more in participants receiving the FMD program than in controls, while fat free mass did not change. The anthropometric changes were accompanied by improvement of insulin resistance as reflected by the Matsuda Index. Average changes in blood pressure and plasma lipid profiles did not differ between groups, except for a slightly larger increase of HDL-cholesterol in FMD users.

The potentially confounding effects of medication on HbA1c levels was accounted for by combining the changes in HbA1c and glucose-lowering medication. For average effects, this was achieved by correcting HbA1c levels for the medication effect score [19]. The MES is a metric that converts an individual’s doses and types of glucose-lowering medications into a summed common metric and illustrates its average HbA1c-lowering potential [26]. Moreover, to comprehensively qualify the status of glycaemic control of individual patients, we constructed a categorical outcome measure that combines HbA1c and the use of glucose-lowering medication, for which we coined the term ‘glycaemic management’. Both measures similarly revealed beneficial effects of the FMD program on glycaemic control.

The percentage of participants who benefitted from the FMD program in terms of HbA1c reduction, diminution of glucose-lowering medication or glycaemic management appeared somewhat higher in the per protocol than in the intention-to-treat analysis. However, the differences between analyses were small, suggesting that less (frequent) dietary intervention may be sufficient to attain guideline goals in a significant proportion of patients. Indeed, further research should aim to define the minimal frequency of FMD cycles required for optimal effect.

One of the strengths of this study is that it involved routine monitoring and treatment by general practitioners, which adds to the generalizability of the findings to real-life clinical settings. Indeed, the fact that glucose-lowering medication was adapted as usual according to Dutch guidelines for the treatment of type 2 diabetes reinforces the notion that the FMD program will have a similar effect in everyday clinical practice. This approach is likely to yield more realistic and clinically relevant results compared to studies where treatment is tightly controlled according to a study protocol.

A limitation of our study concerns the exclusion of patients who used glucose-lowering medication other than metformin. We did so because reduction of caloric intake increases the risk of hypoglycaemia in people using sulphonylurea derivatives or insulin (which were first choice second and third line of (drug) treatment respectively in Dutch guidelines at the time the study started). Therefore, prescription of the FMD program to patients taking these drugs requires more intense surveillance. In a recent trial examining the same dietary intervention, insulin dose was more than halved and all other glucose-lowering drugs were discontinued during FMD, and patients were required to self-monitor blood glucose concentrations at least 4 times daily [27]. In this setting, the FMD program appeared safe, but it was applied to a limited number of patients. Thus, further research is necessary to determine how the FMD program can be safely applied in patients who use glucose-lowering medication other than metformin. Furthermore, missing outcome data in the intention-to-treat analysis may have caused selection bias, although missing data were probably distributed randomly among study groups, since we strongly encouraged people to adhere to (other) protocol instructions even if they discontinued the (dietary) intervention.

In general, the diet program was well tolerated, as illustrated by the similar number of (mild to moderately severe) adverse events and drop-out rates in the FMD and control groups. However, it is important to note that a variety of (minor) complaints were reported during phone calls (meant to promote adherence) at the time participants used the diet, which made five participants discontinue the FMD. It seems prudent to warn people that transient signs of calorie deficit (fatigue, dizziness, headache) may occur during FMD periods. Despite these issues, the majority of participants stayed motivated and complied with the program. This indicates that most individuals will eventually be able to sustainably integrate an FMD program in regular care, which is important as the treatment of type 2 diabetes requires lifelong adaptation of dietary habits.

The results of three previous studies are in line with our findings. Three 5-day cycles of similar composition and timing as used in our trial improved (average) anthropometric measures and metabolic control particularly in obese people with metabolic anomalies at baseline [15], as well as in people with type 2 diabetes [28]. Six cycles improved markers of metabolic control in the FMD group but not in a group with similarly timed cycles of a Mediterranean diet [27] in people with type 2 diabetes. The lack of effect on average HbA1c levels in our study may be due to the fact that we included patients whose glucose levels were well controlled at baseline. Many studies have shown a strong positive correlation between the average baseline HbA1c concentration and its reduction in response to pharmacological intervention [29]. It is quite conceivable that the same goes for lifestyle interventions.

In conclusion, integration of a monthly FMD program in regular care for people with type 2 diabetes who use metformin only and/or diet alone for glycaemic control reduces the need for glucose-lowering medication, improves HbA1c when adjusted for the medication effect score, and improves anthropometric measures. Moreover, it appears to be safe in routine clinical practice.

## Data Availability

The datasets generated during and/or analysed in the current study are available upon reasonable request. Requests should be sent to the FIT trial corresponding email. All proposals requesting data access will need to specify how the data will be used, and all proposals will need approval of the trial co-investigator team before data release.

## Acknowledgements

We gratefully acknowledge the contribution of all participants, the trial steering committee, the general practice centres, supporting staff and research nurses involved in the trial.

## Authors’ relationships and activities

All authors have completed the ICMJE uniform disclosure form at www.icmje.org/coi_disclosure.pdf (available on request from the corresponding author) and declare: financial support was received from Health∼Holland, Top Sector Life Sciences & Health, the Dutch Diabetes Foundation, and L-Nutra for the project; VL is founder and shareholder of L-Nutra (his shares are destined to the Create Cures Foundation and other charitable and research organizations), owns patents licensed to L-Nutra, receives support for travel expenses from L-Nutra and is on the advisory board of L-Nutra; HL has received consulting fees from Royal Philips and was member of the board of trustees of the SCMR and UEMS section Radiology without payment; no other relationships or activities that could appear to have influenced the submitted work.

## Contribution statement

EB and MS contributed equally to this paper. All authors contributed to the study design. EB and MS conducted the trial. EB and MS accessed and verified the data and performed the data analysis. EB, MS, PP and HP prepared the first draft of the manuscript. All authors participated in data interpretation, critical review, and revision of the manuscript, and had final responsibility for the decision to submit for publication. HP is the guarantor of this work and, as such, had full access to all the data in the study and takes responsibility for the integrity of the data and the accuracy of the data analysis.

## References

1. de Cabo R, Mattson MP (2019) Effects of Intermittent Fasting on Health, Aging, and Disease. New England Journal of Medicine 381(26): 2541–2551. 10.1056/NEJMra1905136

[2] Nencioni A, Caffa I, Cortellino S, Longo VD (2018) Fasting and cancer: molecular mechanisms and clinical application. Nat Rev Cancer 18(11): 707–719. 10.1038/s41568-018-0061-0

3. Di Francesco A, Di Germanio C, Bernier M, de Cabo R (2018) A time to fast. Science 362(6416): 770–775. 10.1126/science.aau2095

[4] Mattison JA, Colman RJ, Beasley TM, et al. (2017) Caloric restriction improves health and survival of rhesus monkeys. Nat Commun 8: 14063. 10.1038/ncomms14063

5. Anson RM, Guo Z, de Cabo R, et al. (2003) Intermittent fasting dissociates beneficial effects of dietary restriction on glucose metabolism and neuronal resistance to injury from calorie intake. Proceedings of the National Academy of Sciences of the United States of America 100(10): 6216–6220. 10.1073/pnas.1035720100

[6] Liu H, Javaheri A, Godar RJ, et al. (2017) Intermittent Fasting Preserves Beta-Cell Mass in Obesity-induced Diabetes via the Autophagy-Lysosome Pathway. Autophagy: 0. 10.1080/15548627.2017.1368596

7. van den Burg EL, van Peet PG, Schoonakker MP, van de Haar DE, Numans ME, Pijl H (2023) Metabolic impact of intermittent energy restriction and periodic fasting in patients with type 2 diabetes: a systematic review. Nutr Rev 81(10): 1329–1350. 10.1093/nutrit/nuad015

[8] Lim EL, Hollingsworth KG, Aribisala BS, Chen MJ, Mathers JC, Taylor R (2011) Reversal of type 2 diabetes: normalisation of beta cell function in association with decreased pancreas and liver triacylglycerol. Diabetologia 54(10): 2506–2514. 10.1007/s00125-011-2204-7

[9] Lean MEJ, Leslie WS, Barnes AC, et al. (2018) Primary care-led weight management for remission of type 2 diabetes (DiRECT): an open-label, cluster-randomised trial. Lancet 391(10120): 541–551. 10.1016/S0140-6736(17)33102-1

[10] Lean MEJ, Leslie WS, Barnes AC, et al. (2019) Durability of a primary care-led weight-management intervention for remission of type 2 diabetes: 2-year results of the DiRECT open-label, cluster-randomised trial. The lancet Diabetes & endocrinology 7(5): 344–355. 10.1016/s2213-8587(19)30068-3

[11] Leibel RL, Rosenbaum M, Hirsch J (1995) Changes in energy expenditure resulting from altered body weight. N Engl J Med 332(10): 621–628. 10.1056/nejm199503093321001

[12] Hall KD, Kahan S (2018) Maintenance of Lost Weight and Long-Term Management of Obesity. Med Clin North Am 102(1): 183–197. 10.1016/j.mcna.2017.08.012

[13] Cheng CW, Villani V, Buono R, et al. (2017) Fasting-Mimicking Diet Promotes Ngn3-Driven β-Cell Regeneration to Reverse Diabetes. Cell 168(5): 775–788.e712. 10.1016/j.cell.2017.01.040

[14] Mishra A, Mirzaei H, Guidi N, et al. (2021) Fasting-mimicking diet prevents high-fat diet effect on cardiometabolic risk and lifespan. Nat Metab 3(10): 1342–1356. 10.1038/s42255-021-00469-6

[15] Wei M, Brandhorst S, Shelehchi M, et al. (2017) Fasting-mimicking diet and markers/risk factors for aging, diabetes, cancer, and cardiovascular disease. Science translational medicine 9(377). 10.1126/scitranslmed.aai8700

[16] Ineen. Transparante ketenzorg 2021, rapportage zorggroepen diabetes mellitus, VRM, COPD en asthma. June, 2022. https://ineen.nl/wp-content/uploads/2022/06/Benchmark-transparante-ketenzorg-2021.pdf (accessed Jul 17, 2022).

17. van den Burg EL, Schoonakker MP, van Peet PG, et al. (2020) Fasting in diabetes treatment (FIT) trial: study protocol for a randomised, controlled, assessor-blinded intervention trial on the effects of intermittent use of a fasting-mimicking diet in patients with type 2 diabetes. BMC Endocr Disord 20(1): 94. 10.1186/s12902-020-00576-7

[18] Nederlandse Huisartsen Genootschap. NHG standaard diabetes mellitus type 2. September, 2018. https://richtlijnen.nhg.org/standaarden/diabetes-mellitus-type-2 (accessed Aug 28, 2019). In:

[19] Alexopoulos AS, Yancy WS, Edelman D, et al. (2021) Clinical associations of an updated medication effect score for measuring diabetes treatment intensity. Chronic Illn 17(4): 451–462. 10.1177/1742395319884096

[20] Matsuda M, DeFronzo RA (1999) Insulin sensitivity indices obtained from oral glucose tolerance testing: comparison with the euglycemic insulin clamp. Diabetes Care 22(9): 1462–1470. 10.2337/diacare.22.9.1462

21. Bergman RN, Ader M, Huecking K, Van Citters G (2002) Accurate assessment of beta-cell function: the hyperbolic correction. Diabetes 51 Suppl 1: S212–220. 10.2337/diabetes.51.2007.s212

[22] Utzschneider KM, Prigeon RL, Faulenbach MV, et al. (2009) Oral disposition index predicts the development of future diabetes above and beyond fasting and 2-h glucose levels. Diabetes Care 32(2): 335–341. 10.2337/dc08-1478

[23] US Department of Health and Human services. Common Terminology Criteria for Adverse Events (CTCAE). Nov 27, 2017. https://ctep.cancer.gov/protocoldevelopment/electronic_applications/docs/ctcae_v5_quick_reference_5X7.pdf (accessed Mar 25, 2019).

[24] Adams TD, Davidson LE, Litwin SE, et al. (2012) Health benefits of gastric bypass surgery after 6 years. Jama 308(11): 1122–1131. 10.1001/2012.jama.11164

[25] Lachin JM (2016) Fallacies of last observation carried forward analyses. Clin Trials 13(2): 161–168. 10.1177/1740774515602688

[26] Cox DJ, Oser T, Moncrief M, Conaway M, McCall A (2021) Long-term follow-up of a randomized clinical trial comparing glycemic excursion minimization (GEM) to weight loss (WL) in the management of type 2 diabetes. BMJ Open Diabetes Res Care 9(2). 10.1136/bmjdrc-2021-002403

27. Sulaj A, Kopf S, von Rauchhaupt E, et al. (2022) Six-Month Periodic Fasting in Patients With Type 2 Diabetes and Diabetic Nephropathy: A Proof-of-Concept Study. J Clin Endocrinol Metab 107(8): 2167–2181. 10.1210/clinem/dgac197

[28] Tang F, Lin X (2020) Effects of Fasting-Mimicking Diet and Specific Meal Replacement Foods on Blood Glucose Control in Patients with Type 2 Diabetes: A Randomized Controlled Trial. Oxid Med Cell Longev 2020: 6615295. 10.1155/2020/6615295

[29] Bloomgarden ZT, Dodis R, Viscoli CM, Holmboe ES, Inzucchi SE (2006) Lower baseline glycemia reduces apparent oral agent glucose-lowering efficacy: a meta-regression analysis. Diabetes Care 29(9): 2137–2139. 10.2337/dc06-1120

