## Supplementary for "Integration of a fasting-mimicking diet program in primary care for type 2 diabetes reduces the need for medication – a 12-month randomised controlled trial"

#### Supplementary figures

**Figure S1. Oral glucose tolerance tests of the FMD group and control group at baseline and at 12 months (intention-to-treat analysis)**

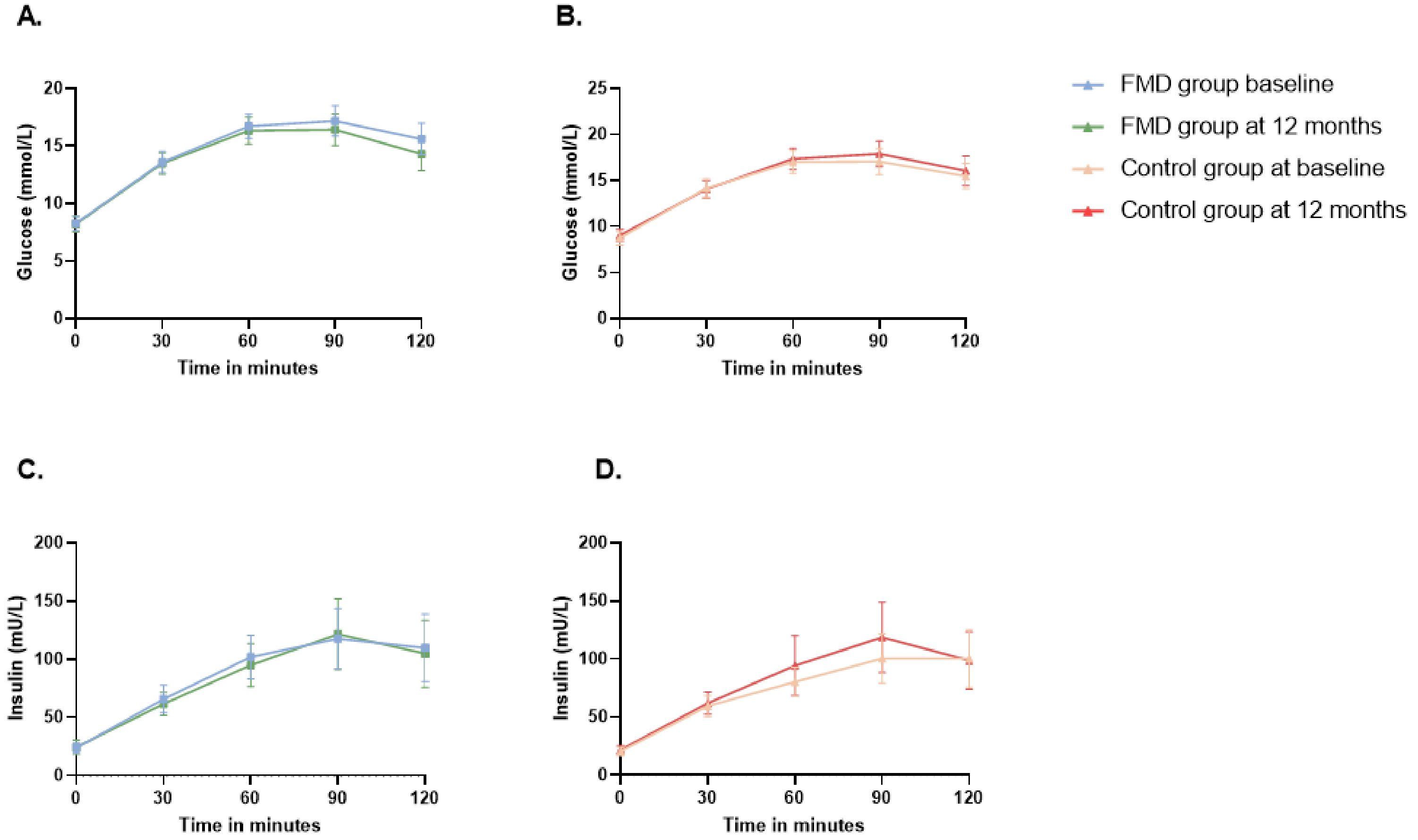

Lines represent the mean levels of glucose or insulin; error bars represent the 95% confidence intervals. FMD n=37, controls n=31.
(A) Glucose measurements of the FMD group. (B) Glucose measurements of the control group. (C) Insulin measurements of the FMD group. (D) Insulin measurements of the control group.
FMD = fasting-mimicking diet.

**Figure S2. HbA1c, glucose-lowering medication and glycaemic management at 12 months (per protocol analyses)**

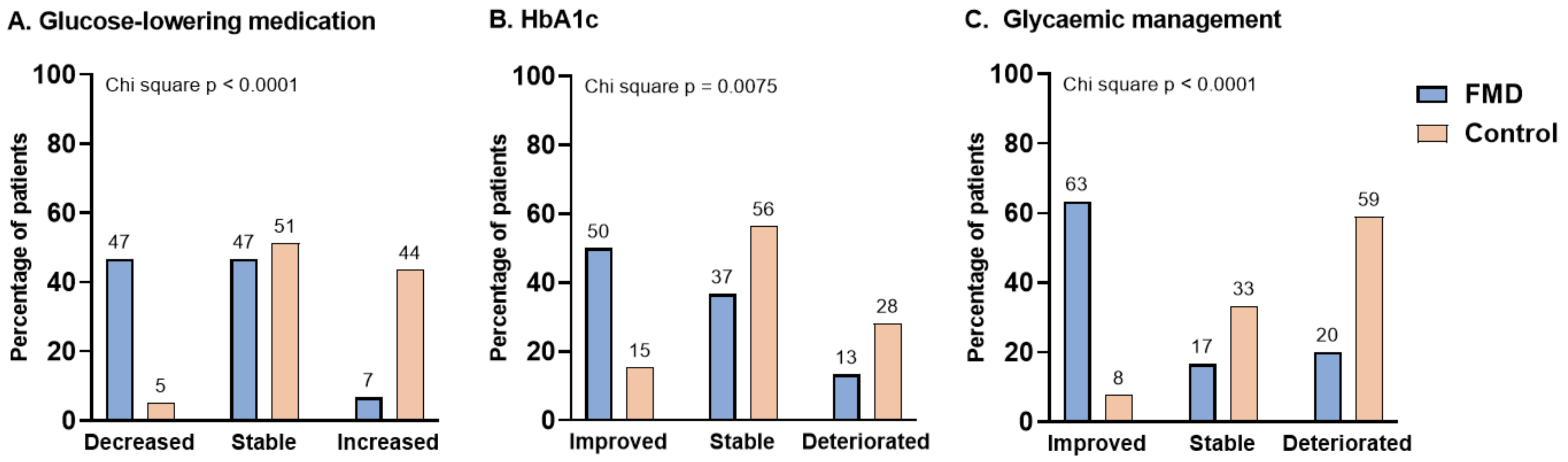

Plotted bars represent percentage of participants. Differences between FMD group and control group were evaluated using the Fisher's exact test or the Chi square test. Number of participants with data available at baseline and 12 months were used for each outcome.

A) Change in glucose-lowering medication, with decrease defined as a lower dose, stable as no change and increased as higher dose or other class of glucose-lowering medication at the end of the study compared to baseline. FMD n=30, controls n=39.

(B) Change in HbA1c, defined as follows; Improved: an HbA1c that is ≥0.5 % (≥5 mmol/mol) lower at the end of the study compared to baseline. Stable: a change in HbA1c of <0.5 % (<5 mmol/mol) at the end of the study compared to baseline. Deteriorated: an HbA1c that is ≥0.5 % (≥5 mmol/mol) higher at the end of the study compared to baseline. FMD n=30, controls n=39.

(C) Glycaemic management, defined as follows; Improved: a lower dose or class of glucose-lowering medication with an HbA1c not more than 0.5 % (5 mmol/mol) higher at the end of the study compared to baseline or; no change in glucose-lowering medication with an HbA1c that is ≥0.5 % (≥5 mmol/mol) lower at the end of the study compared to baseline. Stable: no change in glucose-lowering medication use and a difference in HbA1c of <0.5 % (<5 mmol/mol) at the end of the study compared to baseline. Deteriorated: a higher dose or class of glucose-lowering medication at the end of the study compared to baseline or; an HbA1c that is ≥0.5 % (≥5 mmol/mol) higher at the end of the study compared to baseline with no change in glucose-lowering medication (table 1). FMD n=30, controls n=39.

FMD=fasting-mimicking diet. HbA1c=glycated haemoglobin.

**Figure S3. HbA1c, glucose-lowering medication and glycaemic management at 6 months (intention-to-treat analysis)**

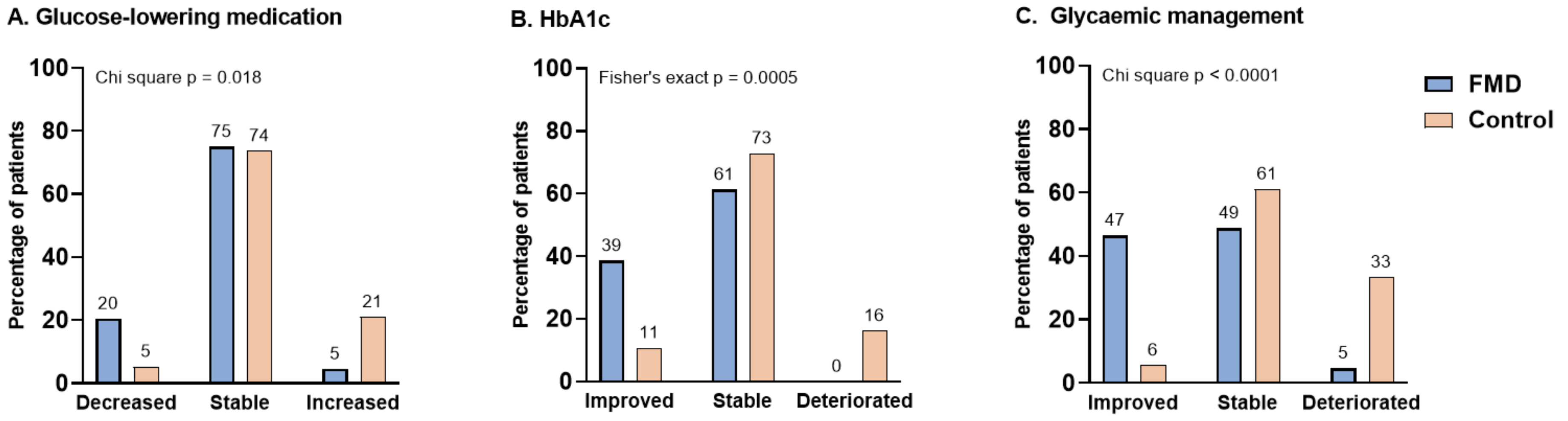

Plotted bars represent percentage of participants. Differences between FMD group and control group were evaluated using the Fisher's exact test or the Chi square test. Number of participants with data available at baseline and 12 months were used for each outcome.

(A) Change in glucose-lowering medication, with decrease defined as a lower dose, stable as no change and increased as higher dose or other class of glucose-lowering medication at the end of the study compared to baseline. FMD n=44, controls n=38

(B) Change in HbA1c, defined as follows; Improved: an HbA1c that is ≥0.5 % (≥5 mmol/mol) lower at the end of the study compared to baseline. Stable: a change in HbA1c of <0.5 % (<5 mmol/mol) at the end of the study compared to baseline. Deteriorated: an HbA1c that is ≥0.5 % (≥5 mmol/mol) higher at the end of the study compared to baseline. FMD n=44, controls n=37

(C) Glycaemic management, defined as follows; Improved: a lower dose or class of glucose-lowering medication with an HbA1c not more than 0.5 % (5 mmol/mol) higher at the end of the study compared to baseline or; no change in glucose-lowering medication with an HbA1c that is ≥0.5 % (≥5 mmol/mol) lower at the end of the study compared to baseline. Stable: no change in glucose-lowering medication use and a difference in HbA1c of <0.5 % (<5 mmol/mol) at the end of the study compared to baseline. Deteriorated: a higher dose or class of glucose-lowering medication at the end of the study compared to baseline or; an HbA1c that is ≥0.5 % (≥5 mmol/mol) higher at the end of the study compared to baseline with no change in glucose-lowering medication. FMD n=43, controls n=36.

FMD=fasting-mimicking diet. HbA1c=glycated haemoglobin.

**Figure S4. Values of HbA1c, MES and selected anthropometrics at baseline, 6 and 12 months for both the FMD and control group (intention-to-treat analysis)**

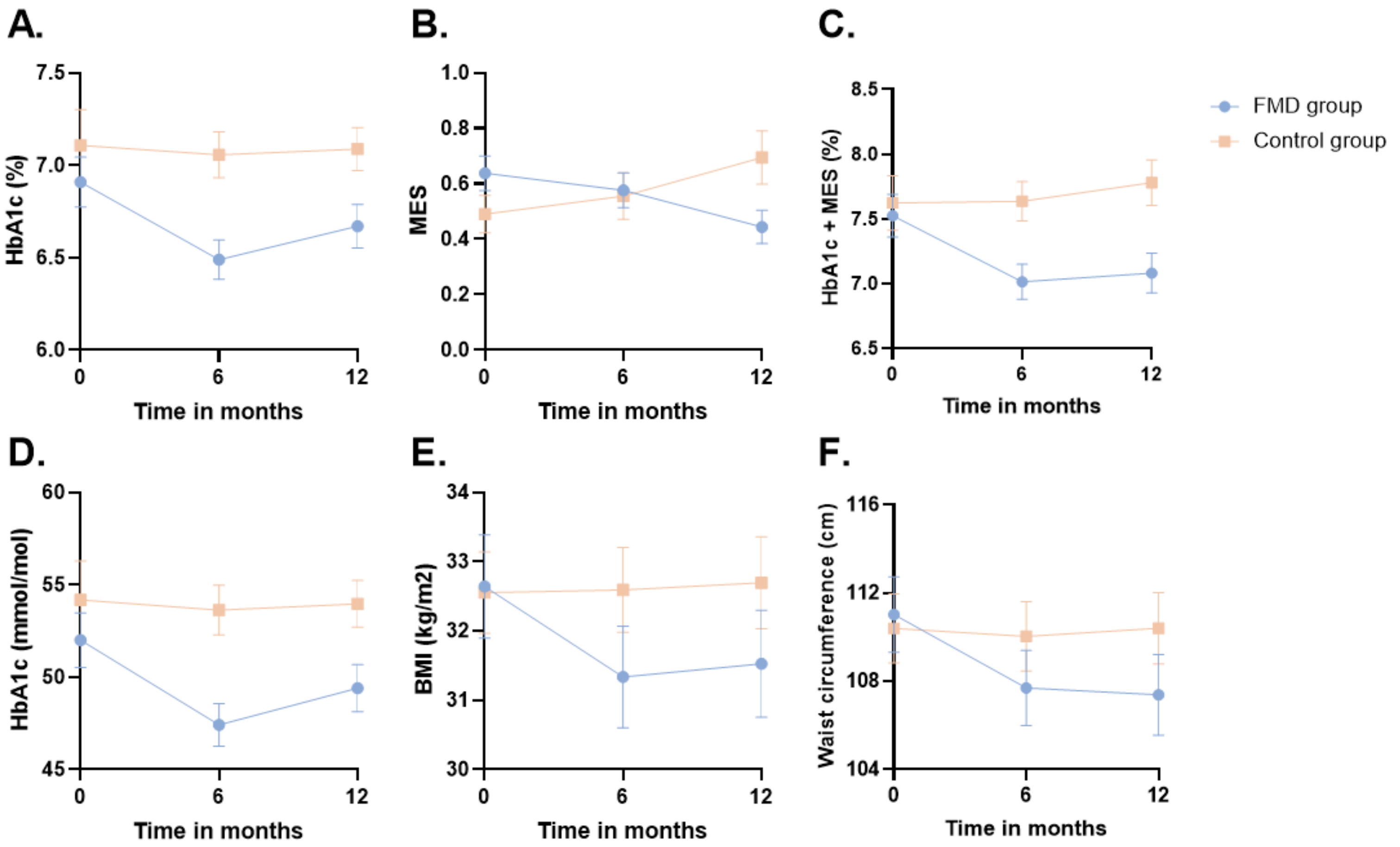

Lines represent the mean values of the FMD group (n=) and the control group (n=) over time. Data was included when available for baseline, 6 and 12 months.
(A) HbA1c (%). FMD n=42, controls n=36. (B) MES. FMD n=40, controls n=36. (C) HbA1c corrected for MES (%). FMD n=40, controls n=35. (D) HbA1c (mmol/mol). FMD n=42, controls n=36. (E) BMI (kg/m$^2$). FMD n=42, controls n=37. (F) Waist circumference (cm). FMD n=42, controls n=37.
BMI=body mass index. FMD=fasting-mimicking diet. HbA1c=glycated haemoglobin. MES=medication effect score.

### Supplementary tables

#### Table S1. Example meal plan of the fasting-mimicking diet for study participants

| | Day 1 | Day 2 | Day 3 | Day 4 | Day 5 |
|---|---|---|---|---|---|
| **Breakfast** | Tea<br>Nut bar<br>Algal Oil capsule | Tea<br>Nut bar | Tea<br>Nut bar | Tea<br>Nut bar | Tea<br>Nut bar<br>Algal Oil capsule |
| **Lunch** | Tomato Soup<br>Olives<br>Kale crackers<br>Vitamin capsule | Tea<br>Mushroom Soup<br>Olives<br>Vitamin capsule | Tea<br>Tomato Soup<br>Kale Crackers<br>Vitamin capsule | Tea<br>Vegetable Soup<br>Olives<br>Vitamin capsule | Tea<br>Tomato Soup<br>Kale Crackers<br>Vitamin capsule |
| **Afternoon** | Tea<br>Nut bar | Tea<br>Olives | Tea | Tea<br>Olives | Tea |
| **Dinner** | Minestrone Soup<br>Choco crisp bar<br>Vitamin capsule | Tea<br>Quinoa Mix Soup<br>Choco crisp bar<br>Vitamin capsule | Tea<br>Minestrone Soup<br>Vitamin capsule | Tea<br>Quinoa Mix Soup<br>Choco crisp bar<br>Vitamin capsule | Tea<br>Minestrone Soup<br>Vitamin capsule |
| **During the day** | | Syrup for water flavouring | Syrup for water flavouring | Syrup for water flavouring | Syrup for water flavouring |

#### Table S2a. Changes over time in HbA1c, MES and HbA1c adjusted for MES as estimated from linear mixed models (intention-to-treat analysis)

| | Estimated effect (95% CI) | p-value |
|---|---|---|
| **HbA1c (mmol/mol)** | | |
| 6 months | -4.39 (-7.71 to -1.07) | 0.01 |
| 12 months | -2.62 (-5.92 to 0.68) | 0.12 |
| **MES** | | |
| 6 months | -0.09 (-0.22 to 0.04) | 0.18 |
| 12 months | -0.36 (-0.49 to -0.23) | <0.0001 |
| **HbA1c, MES corrected (%)** | | |
| 6 months | -0.49 (-0.81 to -0.18) | 0.0025 |
| 12 months | -0.57 (-0.88 to -0.26) | 0.0005 |

Linear mixed models were computed with time, intervention and time*intervention interaction as fixed effects, and individual patients as random effect.

Available HbA1c data: FMD n=49 and control n=43 at baseline, FMD n=44 and control n=37 at 6 months, and FMD n=43 and control n=39 at 12 months.
Available MES data: FMD n=50 and control n=44 at baseline, FMD n=44 and control n=38 at 6 months, and FMD n=42 and control n=39 at 12 months.
Available HbA1c corrected for MES data: FMD n=49 and control n=43 at baseline, FMD n=43 and control n=36, and FMD n=42 and control n=39 at 12 months.

CI=confidence interval. FMD=fasting-mimicking diet. HbA1c=glycated hemoglobin. MES = medication effect score.

#### Table S2b. Post-hoc analysis evaluating the interaction between weight and intervention group (linear mixed models, intention-to-treat analysis)

| | Estimated effect (95% CI) | p-value |
|---|---|---|
| **HbA1c (mmol/mol)** | | |
| weight * intervention | 0.13 (-0.08 to 0.34) | 0.23 |
| **MES** | | |
| weight * intervention | 0.006 (-0.005 to 0.017) | 0.29 |
| **HbA1c, MES corrected (%)** | | |
| weight * intervention | 0.002 (-0.004 to 0.046) | 0.11 |

To the linear mixed models in table S2a, the interaction between weight and intervention group was added as a fixed effect.

Available data and abbreviations: see table S2a.

#### Table S3. Average change of anthropometrics and plasma metabolic profiles from baseline to 12 months in FMD and control group (per protocol analyses)

| | n* | Baseline | | 12 months | | Change | | Intervention effect | |
|---|---|---|---|---|---|---|---|---|---|
| | | Mean | SD | Mean | SD | Mean | SD | **Estimate (95% CI)** | **Between group p-value** |
| **Primary outcomes** | | | | | | | | | |
| HbA1c (mmol/mol) | | | | | | | | -3,5 (-7,9 to 1,0) | 0,12 |
| FMD | 30 | 51,0 | 8,9 | 47,5 | 8,0 | -3,5 | 8,6 | | |
| Control | 39 | 53,9 | 12,4 | 53,8 | 7,6 | 0,0 | 9,6 | | |
| HbA1c (%) | | | | | | | | -0.3 (-0.7 to 0.1) | 0,12 |
| FMD | 30 | 6,8 | 0,8 | 6,5 | 0,7 | -0,3 | 0,8 | | |
| Control | 39 | 7,1 | 1,1 | 7,1 | 0,7 | 0,0 | 0,9 | | |
| MES | | | | | | | | -0.4 (-0.6 to -0.2) | <0,0001 |
| FMD | 29 | 0,6 | 0,4 | 0,4 | 0,3 | -0,2 | 0,4 | | |
| Control | 39 | 0,5 | 0,4 | 0,7 | 0,6 | 0,2 | 0,4 | | |
| HbA1c (%) corrected with MES | | | | | | | | -0.7 (-0.7 to -1.1) | 0,0011 |
| FMD | 29 | 7,4 | 0,9 | 6,9 | 0,9 | -0,5 | 0,9 | | |
| Control | 39 | 7,6 | 1,2 | 7,8 | 1,0 | 0,2 | 0,8 | | |
| **Secondary outcomes** | | | | | | | | | |
| *Laboratory measurements* | | | | | | | | | |
| Fasting glucose (mmol/L) | | | | | | | | -0,4 (-1,3 to 0,6) | 0,45 |
| FMD | 29 | 8,2 | 1,7 | 8,1 | 1,8 | -0,1 | 1,7 | | |
| Control | 38 | 8,8 | 1,9 | 9,0 | 1,8 | 0,3 | 2,0 | | |
| Fasting insulin (mU/L) | | | | | | | | -1,9 (-6,9 to 3,1) | 0,45 |
| FMD | 28 | 23,4 | 11,2 | 23,7 | 18,7 | 0,3 | 10,5 | | |
| Control | 38 | 21,1 | 10,6 | 23,3 | 11,9 | 2,2 | 9,8 | | |
| Cholesterol (mmol/L) | | | | | | | | 0,1 (-0,2 to 0,4) | 0,56 |
| FMD | 30 | 4,8 | 1,0 | 4,9 | 1,0 | 0,1 | 0,6 | | |
| Control | 38 | 4,8 | 1,0 | 4,8 | 1,2 | 0,0 | 0,6 | | |
| LDL Cholesterol (mmol/L) | | | | | | | | 0,0 (-0,2 to 0,3) | 0,69 |
| FMD | 29 | 2,7 | 0,9 | 2,8 | 0,9 | 0,0 | 0,4 | | |
| Control | 38 | 2,7 | 0,8 | 2,7 | 1,0 | 0,0 | 0,5 | | |
| HDL Cholesterol (mmol/L) | | | | | | | | 0,1 (0,1 to 0,2) | 0,0011 |
| FMD | 30 | 1,2 | 0,2 | 1,3 | 0,3 | 0,1 | 0,2 | | |
| Control | 39 | 1,3 | 0,3 | 1,3 | 0,3 | 0,0 | 0,2 | | |
| Cholesterol/HDL ratio | | | | | | | | -0,3 (-0,6 to 0,1) | 0,10 |
| FMD | 30 | 4,1 | 1,2 | 3,9 | 1,1 | -0,3 | 0,8 | | |
| Control | 38 | 3,7 | 1,0 | 3,7 | 0,9 | 0,0 | 0,5 | | |
| Triglycerides (mmol/L) | | | | | | | | -0,2 (-0,6 to 0,1) | 0,20 |
| FMD | 30 | 1,9 | 0,9 | 1,8 | 0,8 | -0,2 | 0,9 | | |
| Control | 38 | 1,7 | 0,7 | 1,8 | 0,8 | 0,0 | 0,6 | | |
| High sensitive CRP | | | | | | | | -0,1 (-1,4 to 1,3) | 0,94 |
| FMD | 30 | 2,7 | 2,8 | 2,0 | 2,1 | -0,8 | 1,3 | | |
| Control | 39 | 3,4 | 3,6 | 2,7 | 2,3 | -0,7 | 3,5 | | |
| *Anthropometrics* | | | | | | | | | |
| Weight (kg) | | | | | | | | -5,0 (-7,1 to -2,8) | <0,0001 |
| FMD | 30 | 99,1 | 10,5 | 94,6 | 10,4 | -4,6 | 5,7 | | |
| Control | 39 | 99,0 | 14,8 | 99,4 | 15,2 | 0,4 | 3,1 | | |
| BMI (kg/m2) | | | | | | | | -1,7 (-2,4 to -1,0) | <0,0001 |
| FMD | 30 | 32,9 | 4,7 | 31,5 | 4,8 | -1,5 | 1,9 | | |
| Control | 39 | 32,5 | 3,5 | 32,6 | 3,9 | 0,2 | 1,0 | | |
| Waist circumference (cm) | | | | | | | | -4,8 (-7,1 to -2,4) | 0,0001 |
| FMD | 30 | 111,3 | 9,7 | 106,8 | 10,3 | -4,5 | 5,6 | | |
| Control | 39 | 110,3 | 9,3 | 110,6 | 9,7 | 0,2 | 4,2 | | |
| Body fat (%) | | | | | | | | -2,7 (-4,1 to -1,3) | 0,0004 |
| FMD | 30 | 37,6 | 8,2 | 35,6 | 9,0 | -2,1 | 3,8 | | |
| Control | 39 | 37,3 | 7,1 | 37,9 | 7,5 | 0,6 | 2,1 | | |
| Fat free mass (kg) | | | | | | | | -0,5 (-1,4 to 0,3) | 0,22 |
| FMD | 30 | 61,5 | 7,9 | 60,5 | 7,8 | -1,0 | 1,9 | | |
| Control | 39 | 62,2 | 12,1 | 61,8 | 11,9 | -0,4 | 1,7 | | |
| Systolic blood pressure (mmHg) | | | | | | | | 0,3 (-6,7 to 7,3) | 0,92 |
| FMD | 30 | 141,3 | 19,3 | 139,4 | 18,2 | -1,8 | 15,7 | | |
| Control | 39 | 141,1 | 15,3 | 139,0 | 14,5 | -2,2 | 13,3 | | |
| Diastolic blood pressure (mmHg) | | | | | | | | -0,7 (-4,0 to 2,7) | 0,68 |
| FMD | 30 | 84,2 | 6,9 | 81,5 | 6,1 | -2,7 | 6,7 | | |
| Control | 39 | 84,0 | 7,9 | 82,1 | 7,0 | -2,0 | 7,1 | | |

*Number of participants with data available at baseline and 6 months for each outcome. BMI=body mass index. CI=confidence interval. CRP=c-reactive protein. FMD=fasting-mimicking diet. HbA1c=glycated haemoglobin.

#### Table S4. Average change of anthropometrics and plasma metabolic profiles from baseline to 6 months in FMD and control group (intention-to-treat analysis)

| | | Baseline | | 6 months | | Change | | Intervention effect | |
|---|---|---|---|---|---|---|---|---|---|
| | **n*** | Mean | SD | Mean | SD | Mean | SD | **Estimate (95% CI)** | **Between group p-value** |
| **Primary outcomes** | | | | | | | | | |
| HbA1c (mmol/mol) | | | | | | | | -4.0 (-7.8 to -0.3) | 0.034 |
| FMD | 44 | 52.0 | 9.4 | 47.3 | 7.4 | -4.6 | 7.0 | | |
| Control | 37 | 54.4 | 12.6 | 53.8 | 8.1 | -0.6 | 9.3 | | |
| HbA1c (%) | | | | | | | | -0,4 (-0,7 to 0,0) | 0,033 |
| FMD | 44 | 6,9 | 0,9 | 6,5 | 0,7 | -0,4 | 0,6 | | |
| Control | 37 | 7,1 | 1,2 | 7,1 | 0,7 | -0,1 | 0,9 | | |
| MES | | | | | | | | -0,1 (-0,2 to 0,0) | 0,084 |
| FMD | 44 | 0,6 | 0,4 | 0,6 | 0,4 | -0,1 | 0,2 | | |
| Control | 38 | 0,5 | 0,4 | 0,5 | 0,5 | 0,0 | 0,3 | | |
| HbA1c (%) corrected with MES | | | | | | | | -0,5 (-0,8 to -0,1) | 0,0054 |
| FMD | 43 | 7,5 | 1,0 | 7,0 | 0,9 | -0,5 | 0,7 | | |
| Control | 36 | 7,7 | 1,2 | 7,6 | 0,9 | 0,0 | 0,8 | | |
| **Secondary outcomes** | | | | | | | | | |
| *Laboratory measurements* | | | | | | | | | |
| Fasting glucose (mmol/L) | | | | | | | | -0.7 (-1.4 to -0.1) | 0.034 |
| FMD | 41 | 8.3 | 1.9 | 7.9 | 1.6 | -0.4 | 1.2 | | |
| Control | 34 | 8.8 | 2.0 | 9.2 | 1.8 | 0.3 | 1.6 | | |
| Fasting insulin (mU/L) | | | | | | | | -0.7 (-4.2 to 2.8) | 0.70 |
| FMD | 40 | 22.4 | 11.0 | 22.7 | 15.0 | 0.3 | 8.0 | | |
| Control | 35 | 21.5 | 10.9 | 22.5 | 9.6 | 1.0 | 7.3 | | |
| Cholesterol (mmol/L) | | | | | | | | 0.0 (-0.3 to 0.2) | 0.89 |
| FMD | 44 | 4.6 | 1.0 | 4.7 | 1.0 | 0.1 | 0.6 | | |
| Control | 36 | 4.8 | 0.9 | 4.9 | 1.1 | 0.1 | 0.5 | | |
| LDL Cholesterol (mmol/L) | | | | | | | | 0.0 (-0.2 to 0.2) | 1.00 |
| FMD | 43 | 2.6 | 0.9 | 2.7 | 0.9 | 0.0 | 0.5 | | |
| Control | 36 | 2.7 | 0.8 | 2.8 | 1.0 | 0.0 | 0.5 | | |
| HDL Cholesterol (mmol/L) | | | | | | | | 0.1 (0.0 to 0.1) | 0.027 |
| FMD | 44 | 1.2 | 0.3 | 1.3 | 0.3 | 0.1 | 0.1 | | |
| Control | 37 | 1.3 | 0.3 | 1.3 | 0.3 | 0.0 | 0.1 | | |
| Cholesterol/HDL ratio | | | | | | | | -0.2 (-0.4 to 0.0) | 0.081 |
| FMD | 44 | 4.0 | 1.1 | 3.8 | 1.0 | -0.2 | 0.5 | | |
| Control | 36 | 3.8 | 0.9 | 3.8 | 0.9 | 0.0 | 0.5 | | |
| Triglycerides (mmol/L) | | | | | | | | -0.2 (-0.5 to 0.1) | 0.22 |
| FMD | 44 | 1.8 | 0.8 | 1.7 | 0.7 | -0.1 | 0.6 | | |
| Control | 36 | 1.8 | 0.7 | 1.8 | 0.9 | 0.1 | 0.6 | | |
| High sensitive CRP | | | | | | | | 1.2 (-0.5 to 2.8) | 0.17 |
| FMD | 44 | 2.6 | 2.6 | 3.0 | 4.2 | 0.3 | 4.2 | | |
| Control | 37 | 3.5 | 3.7 | 2.6 | 2.1 | -0.9 | 3.2 | | |
| *Anthropometrics* | | | | | | | | | |
| Weight (kg) | | | | | | | | -3.8 (-5.3 to -2.3) | <0.0001 |
| FMD | 44 | 99.4 | 14.4 | 95.5 | 14.9 | -3.8 | 3.9 | | |
| Control | 38 | 98.7 | 15.0 | 98.6 | 15.0 | 0.0 | 2.7 | | |
| BMI (kg/m2) | | | | | | | | -1.3 (-1.8 to -0.8) | <0.0001 |
| FMD | 44 | 32.8 | 4.8 | 31.5 | 4.8 | -1.3 | 1.3 | | |
| Control | 38 | 32.5 | 3.6 | 32.5 | 3.7 | 0.0 | 0.9 | | |
| Waist circumference (cm) | | | | | | | | -2.7 (-4.6 to -0.7) | 0.0086 |
| FMD | 44 | 111.2 | 11.1 | 108.0 | 11.3 | -3.3 | 4.8 | | |
| Control | 38 | 110.4 | 9.4 | 109.8 | 9.5 | -0.6 | 3.9 | | |
| Body fat (%) | | | | | | | | -1.9 (-3.2 to -0.7) | <0.0029 |
| FMD | 44 | 37.9 | 8.0 | 36.3 | 7.9 | -1.6 | 3.3 | | |
| Control | 38 | 37.4 | 7.2 | 37.7 | 7.5 | 0.3 | 2.0 | | |
| Fat free mass (kg) | | | | | | | | -0.6 (-1.6 to 0.3) | 0.19 |
| FMD | 44 | 61.5 | 11.0 | 60.5 | 10.1 | -1.0 | 2.5 | | |
| Control | 38 | 61.8 | 12.3 | 61.4 | 11.8 | -0.4 | 1.7 | | |
| Systolic blood pressure (mmHg) | | | | | | | | 1.2 (-5.3 to 7.8) | 0.71 |
| FMD | 44 | 83.5 | 7.2 | 81.8 | 7.2 | -1.6 | 8.5 | | |
| Control | 38 | 84.1 | 8.0 | 82.7 | 8.4 | -1.4 | 7.7 | | |
| Diastolic blood pressure (mmHg) | | | | | | | | -0.3 (-3.8 to 3.3) | 0.89 |
| FMD | 44 | 139.9 | 17.6 | 136.5 | 15.2 | -3.4 | 17.8 | | |
| Control | 38 | 141.3 | 15.3 | 136.6 | 16.3 | -4.6 | 10.5 | | |

*Number of participants with data available at baseline and 6 months for each outcome. BMI=body mass index. CI=confidence interval. CRP=c-reactive protein. FMD=fasting-mimicking diet. HbA1c=glycated haemoglobin.

#### Table S5. Adverse events

| | Reported at regular appointments (6 months and 12 months) | | In FMD period |
|---|---|---|---|
| **Symptom** | **FMD** | **Control** | |
| Bloating | 1 | ·· | 2 |
| Concentration impairment | ·· | ·· | 2 |
| Constipation | 1 | ·· | 2 |
| Diarrhea | 1 | ·· | 9 |
| Dizziness | 1 | 1 | 12 |
| Dyspepsia | ·· | ·· | 2 |
| Fatigue | 1 | 1 | 15 |
| Flatulence | ·· | ·· | 2 |
| Generalized muscle weakness | ·· | ·· | 3 |
| Headache | 1 | 1 | 11 |
| Irritability | ·· | ·· | 1 |
| Muscle cramp | ·· | ·· | 1 |
| Nausea | ·· | ·· | 10 |
| Presyncope | ·· | ·· | 4 |
| Vomiting | ·· | ·· | 4 |
| Other adverse events | 13 | 15 | 57 |
| *of which* | | | |
| Upper respiratory tract symptoms | 1 | 6 | 18 |
| Musculoskeletal | 5 | 8 | 11 |

Number of unique symptoms, there may be several symptoms per participant. At regular appointments (6 months and 12 months), participants were asked for occurrences of adverse events. Between regular appointments, phone calls to participants of the FMD group during the FMD days were made and the mentioned adverse events were reported. Adverse events were reported following the Common Terminology Criteria for Adverse Events (CTCAE) version 5.0.
FMD = fasting-mimicking diet

#### Table S6. Serious adverse events

| Event | Number of occurrences |
|---|---|
| Hospitalization for anaphylactic shock | 1* |
| Hospitalization for cardioversion | 1 |
| Hospitalization for cerebrovascular accident | 1 |
| Hospitalization for decongestive heart failure | 1 |
| Hospitalization for fever due to viral infection | 1 |
| Hospitalization for heart surgery | 1 |
| Hospitalization for hip replacement | 1 |
| Hospitalization for surgery of carcinoma | 1 |

Number of unique events, there may be several symptoms per participant.
*which did not take place during the 5-days of the fasting-mimicking diet
